# Machine Learning Approaches for Electronic Health Records Phenotyping: A Methodical Review

**DOI:** 10.1101/2022.04.23.22274218

**Authors:** Siyue Yang, Paul Varghese, Ellen Stephenson, Karen Tu, Jessica Gronsbell

**Author notes:** **Correspondence to:** Jessica Gronsbell, Postal address: Department of Statistical Sciences, University of Toronto, 700 University Ave., Toronto, ON M5G 1Z5, Canada.

## Abstract

**Objective:** Accurate and rapid phenotyping is a prerequisite to leveraging electronic health records (EHRs) for biomedical research. While early phenotyping relied on rule-based algorithms curated by experts, machine learning (ML) approaches have emerged as an alternative to improve scalability across phenotypes and healthcare settings. This study evaluates ML-based phenotyping with respect to (i) the data sources used, (ii) the phenotypes considered, (iii) the methods applied, and (iv) the reporting and evaluation methods used.

**Materials and Methods:** We searched PubMed and Web of Science for articles published between 2018 and 2022. After screening 850 articles, we recorded 37 variables on 100 studies.

**Results:** Most studies utilized data from a single institution and included information in clinical notes. Although chronic conditions were most commonly considered, ML also enabled characterization of nuanced phenotypes such as social determinants of health. Supervised deep learning was the most popular ML paradigm, while semi-supervised and weakly-supervised learning were applied to expedite algorithm development and unsupervised learning to facilitate phenotype discovery. ML approaches did not uniformly outperform rule-based algorithms, but deep learning offered marginal improvement over traditional ML for many conditions.

**Discussion:** Despite the progress in ML-based phenotyping, most articles focused on binary phenotypes and few articles evaluated external validity or used multi-institution data. Study settings were infrequently reported and analytic code was rarely released.

**Conclusion:** Continued research in ML-based phenotyping is warranted, with emphasis on characterizing nuanced phenotypes, establishing reporting and evaluation standards, and developing methods to accommodate misclassified phenotypes due to algorithm errors in downstream applications.

## BACKGROUND AND SIGNIFICANCE

Electronic health records (EHRs) are a central data source for biomedical research.[1] In recent years, EHR data has been used to support discovery in disease genomics, to enable rapid and more inclusive clinical trial recruitment, and to facilitate epidemiological studies of understudied and emerging diseases.[2–6] EHRs are also positioned to be a key source of data for the development of personalized treatment strategies and generation of real-world evidence.[7,8] A critical aspect of any secondary use of EHR data is phenotyping, the process of identifying patients with a specific phenotype (e.g. the presence or onset time of a clinical condition or characteristic) based on information in their EHR.[9–11] Phenotyping is one of the first steps of an EHR-based application as it is used to both identify and characterize the population of interest.

Generally, the phenotyping consists of 4 steps: (i) data preparation, (ii) algorithm development, (iii) algorithm evaluation, and (iv) application of the algorithm (**Figure 1**). The focus of our article is on the use of machine learning (ML) for algorithm development. Traditionally, phenotypes have been inferred from rule-based algorithms consisting of inclusion and exclusion criteria handcrafted by clinical and informatics experts.[12] However, given the complexity and variation in documentation across phenotypes, providers, and institutions, developing a sufficient set of rules is prohibitively resource-intensive and difficult to scale across conditions and healthcare settings.[13,14] For example, the Electronic Medical Records and Genomics (eMERGE) Network was an early leader in phenotyping in creating a public phenotype library called PheKB. A key finding from this effort was the time intensiveness of rule-based phenotyping, sometimes requiring up to 6-10 months of manual effort depending on the complexity of the condition.[14] Similar findings have been reported by other large research networks such as OHDSI (Observational Health Data Science and Informatics).[10]

**Figure 1.**
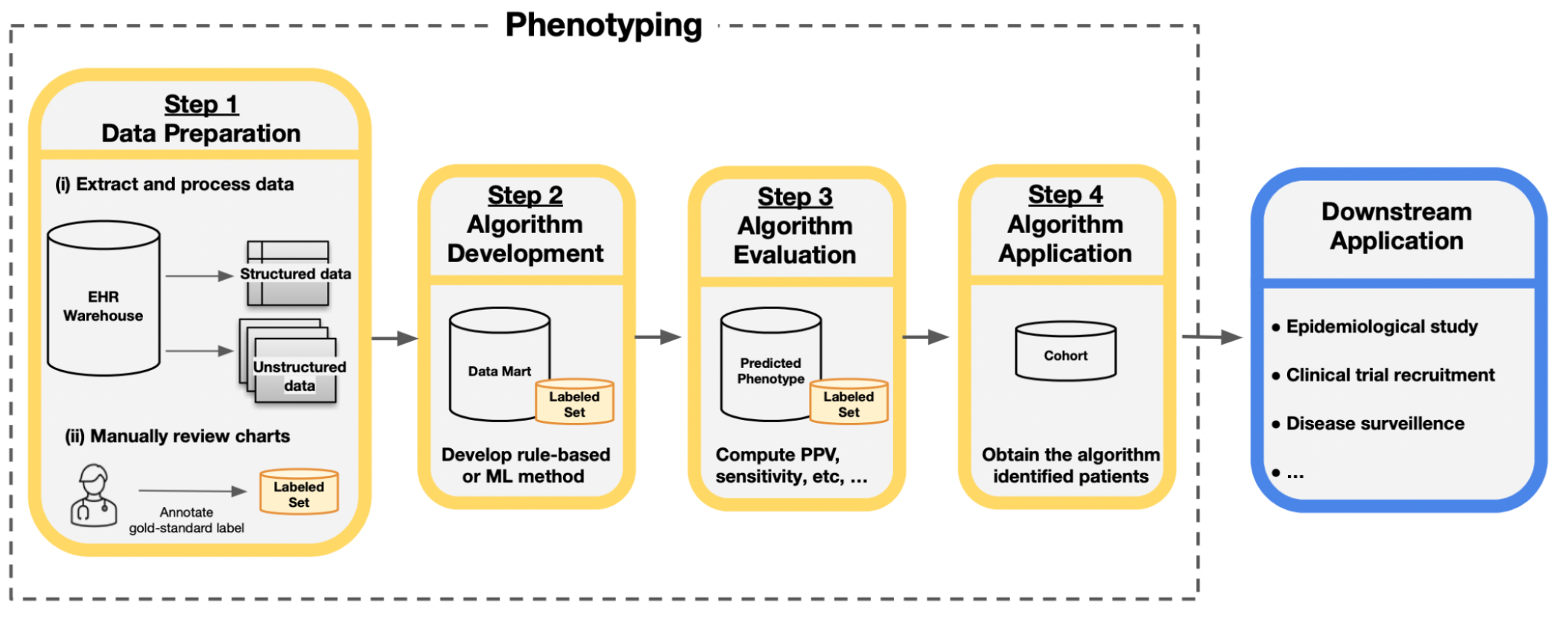
Overview of the phenotyping process. Step 1 involves data preparation which includes (i) extraction and processing of relevant data from records of candidate patients from the data warehouse and (ii) manual review of a subset of charts to obtain gold-standard phenotype labels. Step 2 is the algorithm development phase in which researchers use the data from Step 1, often referred to as the data mart, to develop the phenotyping algorithm with a rule-based or machine learning (ML) method. Step 3 evaluates the accuracy of an algorithm by comparing the assigned phenotype from the algorithm to the gold-standard label, often with estimates of the positive predictive value (PPV), sensitivity, and other accuracy metrics. Step 4 applies the algorithm from Step 2 to obtain the cohort of patients with the phenotype for downstream analysis. The identified cohort can then be used in a variety of downstream applications.

To address this barrier to EHR-based research, there has been increasing interest in phenotyping algorithms derived from ML models.[15,16] In contrast to rule-based approaches, ML methods aggregate multiple sources of information available in patient records in a more automated and generalizable fashion to improve phenotype characterization.[17] While there has been substantial progress in ML approaches designed to make phenotyping more efficient, accurate, and portable in recent years, these advances have yet to be formally synthesized.[18] To the best of our knowledge, 5 articles surveyed EHR-based phenotyping methods through 2018.[11,15–17,21] These articles provide conceptual summaries of rule-based methods and early ML approaches and do not capture advances in semi-supervised, weakly-supervised, and deep learning that were popularized after publication (**Table S1**). Moreover, in light of the wave of EHR-based studies prompted by the pandemic and the increased complexity of ML approaches relative to their rule-based counterparts, there is a pressing need to survey the landscape of phenotyping given its ubiquity in EHR-based applications.[19,20]

## OBJECTIVE

Our work fills this gap in current literature through a methodical review of ML-based phenotyping with respect to (i) the data sources used, (ii) the phenotypes considered, (iii) the methods applied, and (iv) the reporting and evaluation methods used. Based on our analysis of 37 items recorded across 100 selected articles, we also identify areas of future research.

## MATERIALS AND METHODS

### Working definitions

To situate our discussion, key terminology related to EHR data and ML is provided in **Table 1**. We broadly classified a ML method as either (i) supervised, (ii) semi-supervised, (iv) weakly-supervised, or (v) unsupervised according to the model used and the data available for training.[22,23] We further classified each method as deep learning if it is neural network-based and as a traditional ML approach otherwise. Consistent with recent literature, [18] we used an inclusive definition of phenotyping as a procedure that uses EHR data to “assert characterizations about patients.” Our study therefore includes binary phenotypes such as the presence of disease and nuanced phenotypes such as disease severity, disease progression, and social determinants of health (SDOHs). We focused solely on literature using EHRs, defined as longitudinal records of a patient’s interactions with a healthcare institution or system primarily authored by health professionals. We regard our work as a “methodical review” as it does not qualify as a Cochrane style review, but closely adheres to the PRISMA (Preferred Reporting Items for Systematic reviews and Meta-Analyses) guidelines. [24]

**Table 1.** Descriptions of (a) terms used to describe EHR data and (b) ML methods in the context of phenotyping.

| **Term** | **Description** |
| --- | --- |
| Structured data | Data that utilize a controlled vocabulary. Structured data are readily available and searchable in an EHR research database, but often have variable accuracy in characterizing phenotypes. Examples include diagnosis codes, procedure codes, demographics, prescriptions, and laboratory values. |
| Unstructured data | Data that is not organized in a specific manner and requires substantial processing prior to analysis. In the context of phenotyping, the most common form of unstructured data is free-text, such as progress notes, admission and discharge summaries, and radiology reports. Medical images are another form of unstructured data, but were not used in the selected articles. |
| Gold-standard label | The best classification available for phenotype status, most often derived from manual review of patient records by a clinical expert. |
| Silver-standard label | Proxies for the gold-standard phenotype label that are less accurate in characterizing the phenotype, but that can be obtained without time-consuming chart review. Examples include billing codes specific to the phenotype and laboratory values. |
| Feature | Data elements that are potentially predictive of the phenotype and used for algorithm development. Examples include structured data elements such as diagnosis codes and prescriptions as well as information derived from unstructured free-text such as the number of times a phenotype is positively mentioned in a patient's record. |
| Labeled data | Data that contains information on both the gold-standard phenotype labels and the features. |
| Weakly-labeled data | Data that contains information on silver-standard labels and the features. |
| Unlabeled data | Data that contains information on only the features. |

**(b)**
| **ML method** | **Description** | **Motivation for use in phenotyping** |
| --- | --- | --- |
| Supervised learning | Includes methods used to characterize a phenotype with algorithms trained with labeled data. | More automated and potentially more accurate than rule-based methods. |
| Semi-supervised learning | Includes methods used to characterize a phenotype with algorithms trained with both labeled and unlabeled data. | Reduces the amount of labeled data for model training. |
| Weakly-supervised learning | Includes methods used to characterize a phenotype with algorithms trained with weakly-labeled data. | Eliminates the need for labeled data for model training. |
| Unsupervised learning | Includes methods used to identify structure relevant to a phenotype, such as subtypes or clusters of disease progression trajectories, using unlabeled data. | Enables phenotype discovery. |
| Deep learning | A type of ML method that includes methods based on multi-layer neural networks. Can be either supervised, semi-supervised, weakly-supervised, or unsupervised. | Alleviates the need for feature engineering and can yield high accuracy on phenotyping tasks. |
| Traditional machine learning | ML methods that are not constructed based on multi-layer neural networks. | Simpler to implement and interpret than deep learning methods. |

### Search strategy

Due to the broad and evolving definition of phenotyping, early systematic reviews employed a manual review of all full-text articles published in a small number of informatics venues.[12,17] This manual approach was later expanded to a PubMed query [15] using an overly inclusive search designed to capture all articles that (i) used EHR as the primary data source and (ii) utilized ML or natural language processing (NLP) or considered phenotyping. The PubMed query was similarly restricted to a subset of informatics venues in order to target articles focused on phenotyping rather than clinical applications. We followed an analogous strategy, but increased the scope of our search by including Web of Science as we found articles were missed by PubMed. We also added additional strings related to ML.[25]

Specifically, our search of PubMed and Web of Science identified full-text articles that employed ML or NLP or considered phenotyping with EHR data published between January 1, 2018, and April 14, 2022. The range of publication year was specified to not overlap with existing reviews and focused on the same major informatics venues: (1) *Journal of American Medical Informatics Association* (JAMIA), (2) *Journal of Biomedical Informatics* (JBI), (3) *PloS One*, (4) *Proceedings of the American Medical Informatics Association’s Annual Symposium* (AMIA), and (5) *JAMIA Open*.[12,15,16,26,27] The complete search queries are provided in **Table S2**.

### Study selection

Our overall search strategy is depicted in a PRISMA diagram (**Figure 2)**.

**Figure 2.**
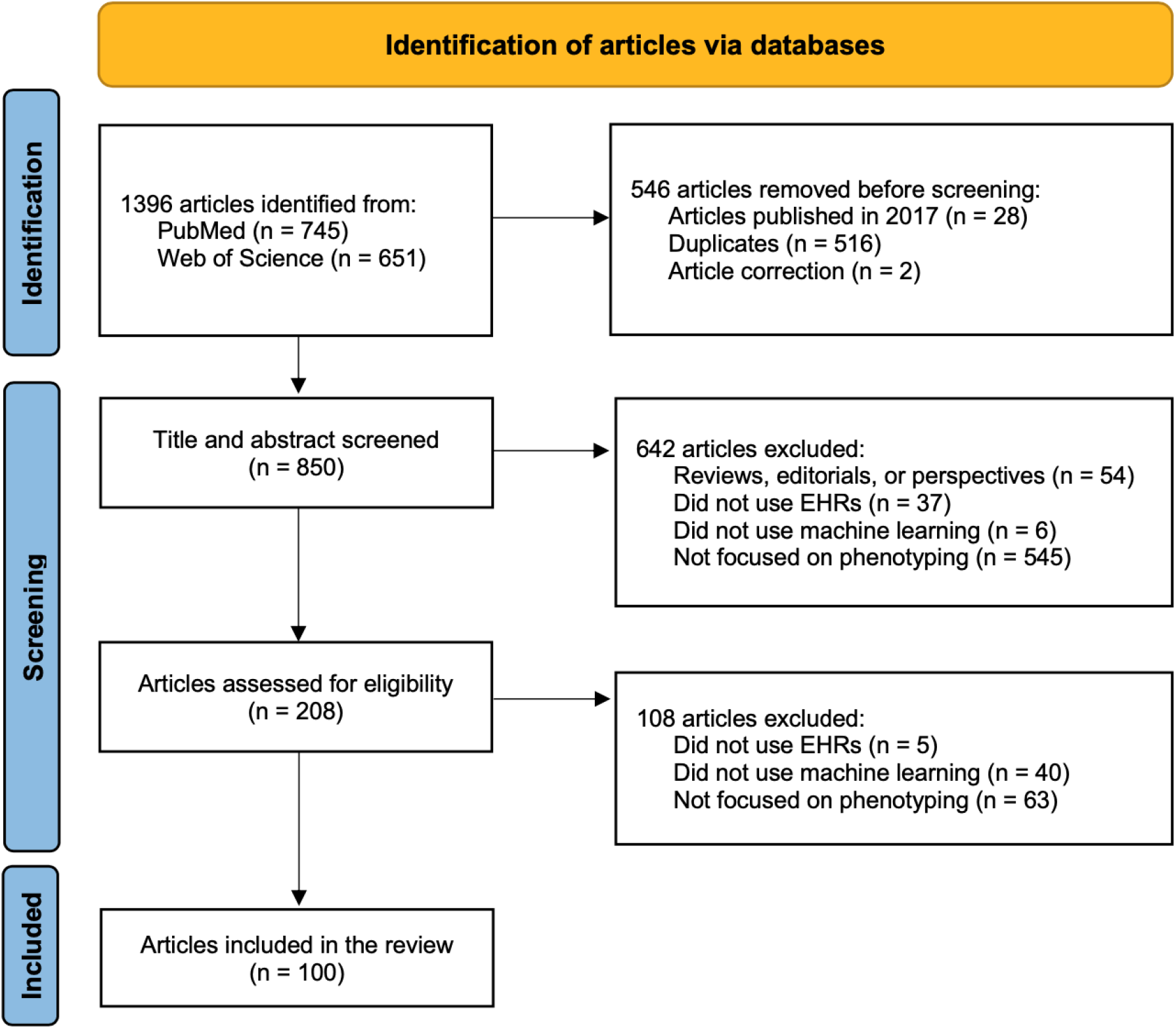
PRISMA diagram for article selection. Only one exclusion reason was chosen for each record during the screening process, although the reasons are not mutually exclusive.

#### Title and abstract screening

After removing duplicates, articles were retrieved and underwent title and abstract screening by two authors (S.Y. and J.G.). A third author (P.V.) resolved disagreements. Articles were excluded if they (i) were reviews, perspectives, or editorials, (ii) did not use EHRs as a primary data source, (iii) did not use ML methods, or (iv) did not consider phenotyping. **Table S3** provides a list of article exclusions.

#### Full-text review

One author (S.Y.) reviewed the full-text articles and another author (J.G.) verified the information from the full-text review when necessary. After excluding papers that did not focus on ML approaches for EHR phenotyping, 100 papers were selected (**Table S4**). During the full-text review, we extracted information on: (i) the data sources used, (ii) the phenotypes considered, (iii) the methods applied, and (iv) the reporting and evaluation methods used. A list of the 37 recorded variables is included in **Table S5**.

## RESULTS

In reviewing the literature, we found that all but two deep learning approaches were supervised (**Figure 3**). We therefore summarize contributions in traditional supervised, deep supervised, semi-supervised, weakly-supervised, and unsupervised learning in the subsequent sections.

**Figure 3.**
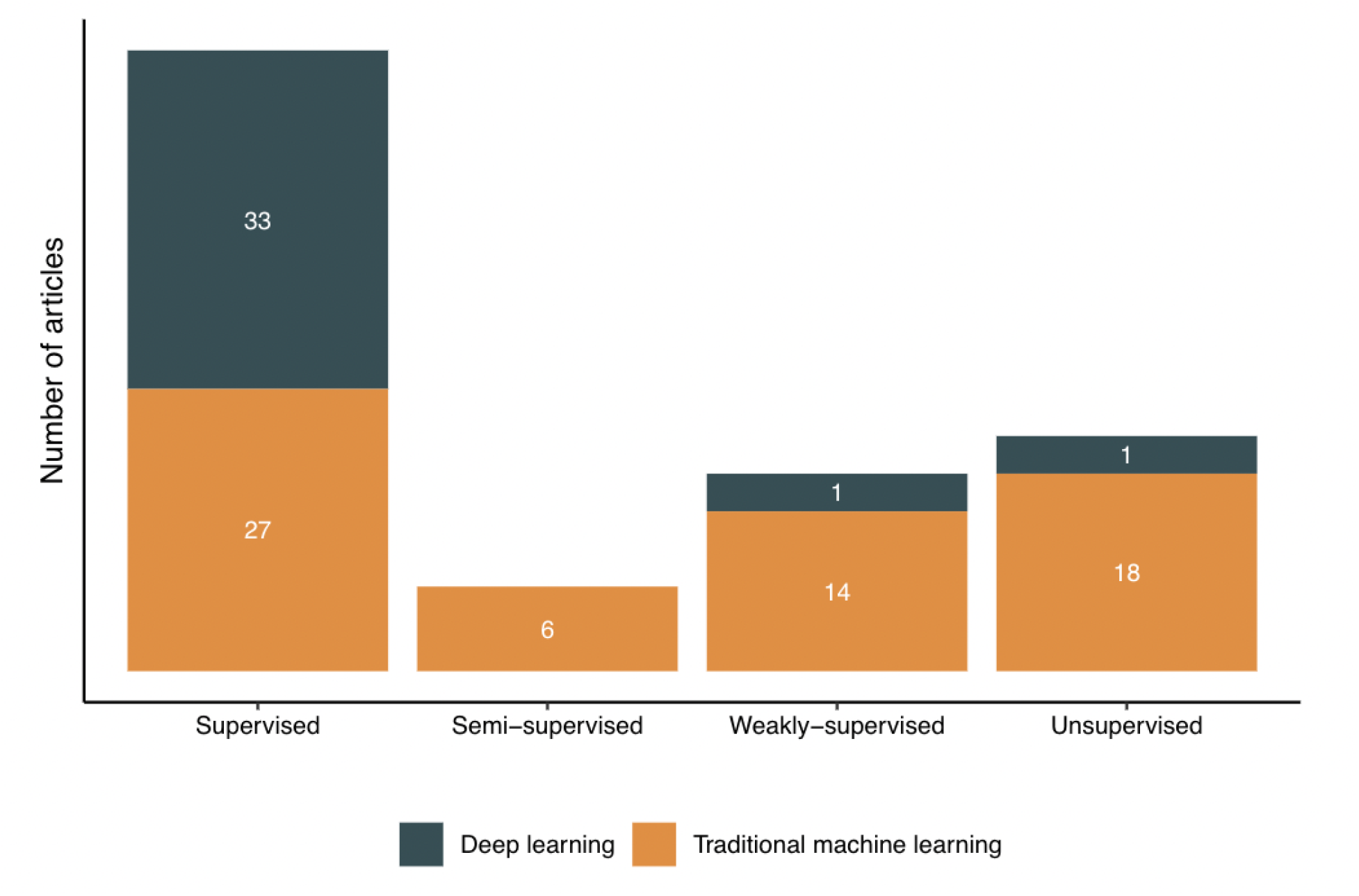
Number of articles that used the various machine learning paradigms.

### Data Sources

63 of the 100 articles relied on EHR data from a single institution, while 8 articles used data from multiple institutions, including research networks such as the OHDSI [28] and eMERGE.[29] The remaining articles leveraged publicly available data from the Medical Information Mart for Intensive Care (MIMIC-III) database and NLP competitions (**Table S6**). A small number of studies utilized additional data sources, including administrative claims [30–36] and registry databases.[37–40] 94 studies were conducted in the US.

With respect to the data types used for developing phenotyping algorithms, 70 of the 100 articles utilized unstructured free-text data and half of these articles also incorporated information from structured data. Unsurprisingly, diagnoses were the most common structured data element and were typically derived from International Classification of Diseases, 9th or 10th Revision (ICD-9/10) billing codes (**Figure 4(a))**. Clinical note types (eg. progress notes, discharge summaries) used for algorithm development were rarely specified (**Figure 4(b))**. However, most articles reported on the NLP software that was used to process free-text. The clinical Text Analysis and Knowledge Extraction System (cTAKEs) was the most popular. Frequently used terminologies and NLP software are detailed in **Table S7** and **S8**, respectively.

**Figure 4.**
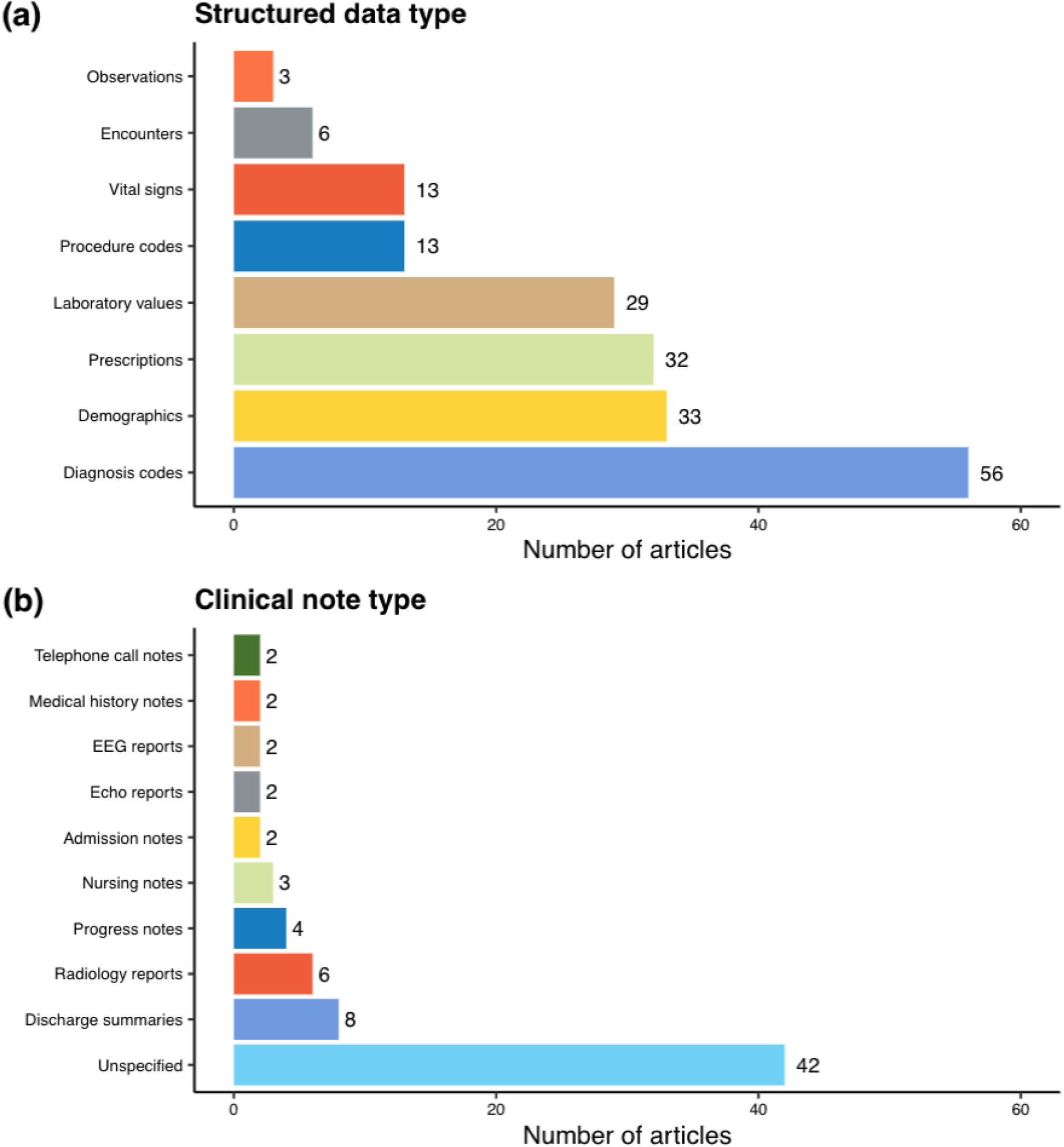
Types of structured data and clinical notes used to develop phenotyping algorithms in the selected articles (excluding articles using competition data). A data type is presented if it is used in more than one article. Encounters include encounter metadata, while medical history notes include both social history and cardiac surgical history.

### Phenotypes

The articles in our study considered 157 phenotypes, with 40 articles focusing on more than one phenotype. Studies using data from NLP competitions focused on adverse drug events [41] and clinical trial eligibility,[42] while studies using MIMIC-III characterized 25 phenotypes seen in the intensive care unit.[43] Outside of the articles using public data, chronic conditions with a large burden on the healthcare system, such as heart diseases and type II diabetes mellitus, were most frequently considered overall. 69 of the 100 articles aimed to identify binary phenotypes (e.g. case/control disease status), while few focused on severity or temporal phenotypes (4 and 11 articles, respectively). Although this finding coincides with previous reviews, there were considerable differences in the top phenotypes across the 5 ML paradigms (**Figure 5**). The phenotypes considered in articles utilizing traditional supervised learning were not identified in previous reviews[12,15] These include phenotypes primarily documented in free-text such as suicidal behavior [44,45] and SDOHs.[30,46–49] Deep supervised learning papers similarly considered SDOHs [50–57] as well as episodic conditions [58–61] and COVID-19.[62,63] The phenotypes considered by articles using semi- or weakly-supervised methods aiming to expedite algorithm development included common, chronic conditions [64–66] that had been previously identified with a rule-based or traditional supervised learning method.[13,67] Most unsupervised methods considered progressive conditions associated with multiple comorbidities or phenotypic heterogeneity such as dementia and chronic kidney disease.[68,69]

**Figure 5.**
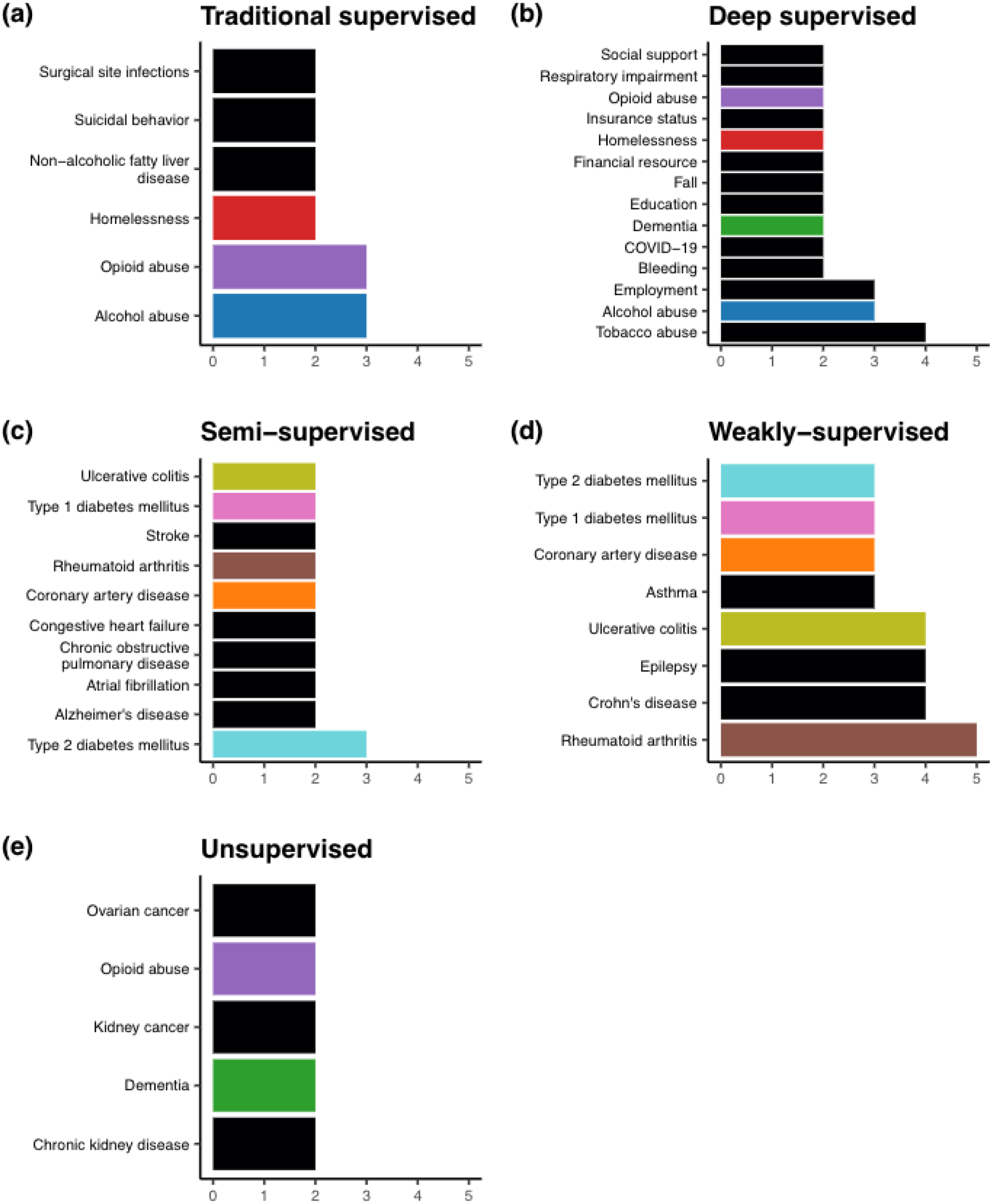
Top phenotypes considered within each machine learning category and the number of articles of each phenotype (excluding articles using competition data sources). Phenotypes are colored if they appear in more than one category.

### ML Methods

#### Traditional supervised learning

60 articles employed supervised learning methods, with 27 articles using traditional models. In contrast to rule-based algorithms, phenotyping algorithms derived from supervised learning are less burdensome to develop as they are learned from the data.[15] Traditional supervised learning is also more amenable to incorporating a greater number of features predictive of the phenotype into the algorithm, such as information in clinical notes.[17] Among the articles using traditional supervised learning, half of them mapped terms in free-text to clinical concepts in the Unified Medical Language System (UMLS) [70] for use in algorithm development. Similar to features derived from structured data elements, the extracted concepts were typically engineered into patient-level features (e.g. total number of positive mentions of a concept in the record) based on the consensus of domain experts.[71] Gold-standard labels for model training were predominantly annotated through manual review of patient records.[72] In some instances, labels were also derived from registry data,[37] laboratory results,[35,36,73] or rule-based algorithms.[47]

The most commonly used methods were random forest, logistic regression, and support vector machine (**Table 2**). A common trend among selected articles was the use of a selective sampling method, such as undersampling or the Synthetic Minority Oversampling Technique (SMOTE), to address class imbalance for rare phenotypes such as surgical site infections and rhabdomyolysis.[31,33,35,37,48,74,75] Several models, including SVM, single-layer perceptron, and logistic regression, were also extended to accommodate federated analysis of distributed EHR data held locally at multiple institutions to identify adverse drug reactions.[33]

**Table 2.** Common methods in each machine learning category. A method is presented if it appeared in more than one article.

| Machine learning type | Method | Number of articles |
| --- | --- | --- |
| Traditional supervised learning | Random forest | 14 |
|  | Logistic regression | 11 |
|  | Support vector machine (SVM) | 11 |
|  | L1 logistic regression | 8 |
|  | Decision trees | 4 |
|  | Extreme gradient boosting (XGBoost) | 4 |
|  | Naive Bayes | 3 |
| Deep supervised learning | Recurrent neural networks (RNNs) and variants | 19 |
|  | Convolutional neural networks (CNNs) and variants | 11 |
|  | BERT and variants | 7 |
|  | Feed-forward neural networks (FFNNs) | 3 |
| Weakly-supervised learning | PheNorm [108] | 3 |
|  | MAP [109] | 2 |
|  | Random forest (with silver-standard labels) | 2 |
| Unsupervised learning | Latent Dirichlet Allocation (LDA) | 5 |
|  | K-means | 4 |
|  | UPGMA (unweighted pair group method with arithmetic mean) hierarchical clustering | 2 |

**Note:** Some papers used more than one method. The table does not include any semi-supervised methods as each article used a distinct method. Semi-supervised methods are presented in Table 3.

#### Deep supervised learning

While traditional supervised learning methods enable the use of free-text in algorithm development, they are limited by their inability to handle raw input data. Deep learning models consist of many processing layers that discover intrinsic patterns within data to alleviate the burden of feature engineering.[76,77] This is particularly valuable in the context of EHR data as models can learn rich representations of the clinical language in free-text.[78] All but 2 articles employing deep supervised learning articles leveraged clinical notes. The articles utilized word embeddings to represent words or clinical concepts as real-valued vectors based on their context.[79] Word embeddings are typically learned from a large corpus in an unsupervised fashion and used as the input layer to a neural network. Common corpora within the selected articles included clinical notes [53,57,63,80–84] as well as external sources such as biomedical publications [56,61,62,85,86] and Wikipedia articles [51,58,87–90] (**Table S9**). Word2vec,[91] Global Vectors (GloVE),[92] and Bidirectional Encoder Representations from Transformers (BERT)[93–96] were the most frequently used methods for training embeddings (**Table S10**).

*Among* neural network architectures, feed-forward networks were only used in 3 studies (**Table S11**)[97] while BERT and variants were frequently used for phenotypes documented in clinical notes such as SDOHs (e.g. education [50,57]) and symptoms (e.g. chest pain,[90] bleeding [58]).

Recurrent neural networks (RNNs), convolutional neural networks (CNNs), and their variants were the most prevalent architectures as they accommodate sequential data in longitudinal patient records and clinical text.[24,76] For instance, the bidirectional long-short term memory (Bi-LSTM), an RNN variant that captures previous and future information in a sequence, was used to characterize phenotypes evolving over time such as dementia [34] and substance abuse.[54] In terms of text-based phenotyping, the Bi-LSTM with a conditional random field layer (Bi-LSTM-CRF) was used to improve identification of adverse drug events.[80,81,88] Similarly, Gehrmann et al. improved text-based phenotyping with a CNN designed to identify phrases relevant to substance abuse, depression, and other chronic conditions with the MIMIC-III phenotype dataset.[55]

#### Semi-supervised learning

Despite its widespread use, supervised learning is difficult to scale due to the time and resources required to obtain gold-standard labeled data.[98] Semi-supervised methods are trained with a large amount of unlabeled data (i.e. unreviewed medical records) and a small amount of labeled data to minimize the burden of chart review.[99] Three types of methods were used in 6 articles utilizing semi-supervised learning (**Table 3)**. The first type performed feature selection using “silver standard labels” that can be automatically extracted from patient records, such as the frequency of phenotype specific diagnostic codes, prior to supervised training.[100,101] For instance, PheCAP processed openly available knowledge sources such as Wikipedia articles to generate a candidate list of related UMLS concepts. An ensemble sparse regression approach using silver-standard labels was then used to identify relevant concepts for supervised learning. PheCAP was used to phenotype over twenty conditions using EHR data from 4 institutions.[100,102] The second type of semi-supervised learning applied self-learning to train a generative model with labeled data to create pseudo-labels for the unlabeled dataset in order to train a traditional supervised model. Self-learning performed on par with supervised learning for 18 phenotypes.[64,65] In contrast, the third type directly incorporated unlabeled data into the algorithm through modification of the loss function.[66,103] For example, a semi-supervised tensor factorization (PSST) approach used the information in unlabeled data to incorporate cannot link constraints into tensor factorization for classification of hypertension and type-2 diabetes.[66] PSST performed similarly to supervised tensor factorization, but with fewer labeled examples.

**Table 3.** Semi-supervised methods used in the selected articles, as well as the phenotypes considered and the size of the labeled and unlabeled datasets.

| Method | Paper | Phenotype(s) | Unlabeled dataset size | Labeled dataset size |
| --- | --- | --- | --- | --- |
| Silver-standard based feature selection | Cade et al.[100] | Sleep apnea | 15,741 | 300 |
|  | Cohen et al.[101] | Acute hepatic porphyria | 22,372 | 200 |
| Self-learning | Estiri et al.[64] | Alzheimer's disease; Atrial fibrillation; Asthma; Bipolar disorder; Breast cancer; Coronary artery disease; Crohn's disease; Congestive heart failure; Chronic obstructive pulmonary disease; Epilepsy; Gout; Hypertension; Rheumatoid arthritis; Schizophrenia; Stroke; Type 1 diabetes mellitus; Type 2 diabetes mellitus; Ulcerative colitis | 5,732 (Average) | 360 (Average) |
|  | Estiri et al.[65] | Alzheimer's disease; Atrial fibrillation; Coronary artery disease; Congestive heart failure; Chronic obstructive pulmonary disease; Rheumatoid arthritis; Stroke; Type 1 diabetes mellitus; Type 2 diabetes mellitus; Ulcerative colitis | 6,000 (Average) | 351 (Average) |
| Modified loss function | Zhang et al.[103] | Aldosteronism | 6,391 | 185 |
|  | Henderson et al.[66] | Resistant hypertension; Type 2 diabetes mellitus | 1,622 | 400 |

#### Weakly-supervised learning

Analogous to semi-supervised learning, the goal of weakly-supervised learning is to expedite the phenotyping process by eliminating the need for gold-standard labeled data. Weakly supervised methods rely on a “silver-standard” label that can be easily extracted from patients records in place of gold standard labels.[104] The silver-standard label is selected based on clinical expertise as a proxy for the phenotype.[104–107] Common silver-standard labels included specific diagnosis codes, lab results, and free-text mentions of the phenotype.[108–110]

Two types of weakly-supervised learning approaches were used in 15 articles (**Table 4)**. The first type assumed the silver-standard label follows a mixture model representing phenotype cases and controls.[108–114] For example, PheNorm utilized Gaussian mixture-models with denoising self-regression for phenotyping 4 chronic conditions.[108] MAP later improved upon PheNorm with an ensemble of mixture models and was validated across 16 phenotypes and two phenome-wide association studies.[40,109] PheVis extended the resolution of PheNorm from patient-level to visit-level by incorporating past medical history information into estimation.[112] The second type of weakly-supervised methods used silver standards to directly train supervised models.[51,105,106,115–119] For instance, APHRODITE employs “noisy label” learning with an anchor feature with a near perfect positive predictive value (PPV), but potentially imperfect sensitivity to train L1-penalized logistic regression models.[115] APHRODITE is available in openly available software for users of the OMOP common data model. Similar approaches have been used to identify phenotypes poorly documented in structured data such as systemic lupus erythematosus.[51,116] In general, weakly-supervised models exhibit similar or improved performance to their rule-based and supervised counterparts (**Figures S1** and **S2**).

**Table 4.** Weakly-supervised methods used in the selected articles, as well as the phenotypes considered and the silver-standard label used.

| Method | Paper | Phenotype(s) | Silver-standard label(s) |  |  |  |
| --- | --- | --- | --- | --- | --- | --- |
|  |  |  | ICD code | SNOMED code | Relevant concept or word in free-text | Other |
| Mixture modeling | PheNorm[108] | Rheumatoid arthritis; Crohn's disease; Ulcerative colitis; Coronary artery disease | ✓ |  | ✓ |  |
|  | PheProb[111] | Rheumatoid arthritis | ✓ |  |  |  |
|  | Multimodel Automated Phenotyping (MAP)[109] | Asthma; Crohn's disease; Ulcerative colitis; Cardiomyopathy; Congestive heart failure; Epilepsy; Juvenile rheumatoid arthritis; Chronic pulmonary heart disease; Type 1 diabetes mellitus; Cardiovascular disease; Inflammatory bowel disease | ✓ |  | ✓ |  |
|  | Geva et al.[40] | Asthma; Bipolar disorder; Schizophrenia; Breast cancer; Chronic obstructive pulmonary disease; Congestive heart failure; Coronary artery disease; Hypertension; Depression; Epilepsy; Multiple sclerosis; Rheumatoid arthritis; Type 1 diabetes mellitus; Type 2 diabetes mellitus; Crohn's disease; Ulcerative colitis | ✓ |  | ✓ |  |
|  | PheMAP [110] | Type 2 diabetes mellitus; Dementia; Hypothyroidism |  |  | ✓ |  |
|  | PheVis [112] | Rheumatoid arthritis; Tuberculosis | ✓ |  | ✓ |  |
|  | Surrogate-guided ensemble latent Dirichlet allocation | Asthma; Breast cancer; Chronic obstructive pulmonary disease; Depression; Epilepsy; |  |  |  | Phenotype probabilities derived from |
|  | (sureLDA)[113] | Hypertension; Schizophrenia; Stroke; Type 1 diabetes mellitus; Obesity |  |  |  | PheNorm |
|  | Ning et al.[114] | Coronary artery disease; Rheumatoid arthritis; Crohn's disease; Ulcerative colitis; Pulmonary hypertension | ✓ |  | ✓ |  |
| Noisy labeling | Automated PHeNotype Routine for Observational Definition, Identification, Training and Evaluation (APHRODITE)[115] | Appendicitis; Type 2 diabetes mellitus; Cataracts; Heart failure; Abdominal aortic aneurysm; Epilepsy; Peripheral arterial disease; Obesity; Glaucoma; Venous thromboembolism |  | ✓ |  |  |
|  | Murray et al.[116] | Systemic lupus erythematosus | ✓ |  |  |  |
|  | Ling et al.[38] | Metastatic breast cancer |  |  | ✓ |  |
|  | Banerjee et al.[119] | Urinary incontinence; Bowel dysfunction |  |  | ✓ |  |
|  | NimbleMiner [118] | Fall |  |  | ✓ |  |
|  | Annapragada et al.[51] | Child physical abuse |  |  | ✓ |  |
|  | Sanyal et al.[117] | Insulin pump failure |  |  | ✓ |  |

#### Unsupervised learning

In contrast to the previously discussed ML approaches, unsupervised learning is used for phenotype discovery, including identification of subphenotypes,[39,74,120–128] co-occurring conditions,[69,129] and disease progression patterns.[68,130–134] Among the 19 articles utilizing unsupervised learning, Latent Dirichlet Allocation (LDA)[69,124,125,127,133] and K-means were the most frequently used methods.[120,121,123,125] LDA was applied to identify the co-occurrence of allergic rhinitis and osteoporosis among patients with kidney disease [69] as well as to capture trends in mental health and end of life care among dementia patients.[133] K-means was used to discover subphenotypes such as patients with different symptoms of acute kidney injury.[120] Model-derived subpopulations were commonly used in downstream prediction tasks.[39,68,121,122,125,131] For example, a SVM was used to identify sepsis using features of subpopulations with distinct dysfunction patterns discovered from a self-organizing map.[128] Only 1 article utilized a deep learning approach, specifically a deep autoencoder to discover subtypes of depression.[132]

### Reporting and Evaluation Methods

As the articles primarily focused on identifying disease cases (excluding unsupervised learning articles), most evaluated algorithm performance with PPV, sensitivity, and/or F-score (70/81 articles reported at least one of these metrics; **Table S12**). The area under the ROC curve (AUROC) was also reported as an overall summary of discriminative performance (42/81 articles), while calibration was rarely assessed (5/81 articles). Additionally, several studies linked EHR data to administrative claims [30–36] or registry databases [37–40] to validate algorithm accuracy. Biorepositories were also used to demonstrate the validity of a derived phenotype in replicating a genetic association study.[109–111,135] Only 5 studies performed external validation or evaluated algorithmic fairness.[36,40,52,61,136] We also found limited reporting of the data descriptors necessary to assess the feasibility of implementing an algorithm in a new setting. Patient demographics were only reported in 38 of 71 papers using private data sources and only 20 articles released their analytic code. A majority of these articles used complex deep learning models (9 articles) and/or free-text data (9 articles).

With respect to performance comparisons, 21 articles compared an ML approach to a rule-based method (**Table S13**). Traditional ML was used in 10 of these articles and outperformed rule-based algorithms in 8 articles with respect to PPV, sensitivity, or both **(Figure S3)**. 2 supervised deep learning models were compared to rules, with a Bi-LSTM performing similarly to a rule-based approach for substance abuse [54] and a bidirectional gated recurrent unit model significantly decreasing performance in identifying insulin rejection.[137] 20 articles also provided comparisons between deep learning and traditional baselines (**Table S14**). Deep learning outperformed traditional ML across all reported accuracy metrics for 18 of 33 phenotypes considered (**Figure S4(a)**). Deep learning improved sensitivity with a corresponding decrease in PPV or vice-versa (**Figure S4(b-c)**) for the remaining phenotypes, with the exception of one study demonstrating that elastic net logistic regression outperformed a RNN for phenotyping fall risk (**Figure S4(d**)).[61] It is important to note that a meaningful gain in accuracy must be interpreted in the context of the use case of the algorithm and the target metric of performance. Moreover, improvements in accuracy must be weighed against additional challenges brought on by deep learning, including data demands, decreased interpretability, and limited generalizability over time and across healthcare settings.[72,138–140]

## DISCUSSION

This review highlights the substantial ongoing work in ML-based phenotyping. A broad range of phenotypes have been considered and the use of unstructured information in clinical notes is widespread. While ML approaches did not uniformly outperform rule-based methods, deep learning provided marginal improvement over traditional baselines. Moreover, semi-supervised and weakly-supervised learning have expedited the phenotyping process while unsupervised learning has been effective for phenotype discovery. Progress withstanding, most articles focused on binary phenotypes and few studies evaluated external validity or used multi-institution data. Study settings were infrequently reported and analytic code was rarely released. Future work is warranted in “deep phenotyping”, reporting and evaluation standards, and methods to accommodate misclassified phenotypes due to algorithm errors in downstream applications.

### Deep Phenotyping

“Deep phenotyping” moves beyond binary identification to characterization of nuanced phenotypes, such as the timing or severity of a condition, using advanced methods leveraging interoperable and multimodal data types.[19, 122,141,142] From a methodological viewpoint, studies of nuanced phenotypes will face similar, but more substantial challenges in obtaining gold-standard labeled data. Further work in semi- and weakly-supervised deep learning methods is necessary.[143,144] Moreover, given the privacy constraints associated with EHRs and other health data sources, leveraging interoperable and multimodal data calls for advancements in federated learning methods that can accommodate distributed data sources stored locally across institutions. [145]

### Reporting & Evaluation Standards

Research networks, such as eMERGE, have long advocated for transparent and reusable phenotype definitions. Most recently, in response to the wave of COVID-19 studies, Kohane et al. proposed a checklist for evaluating the quality of EHR-based studies, emphasizing phenotypic transparency as a key concern.[146] However, we found most articles did not release necessary details for complete evaluation of an approach or implementation in other settings. As a step towards reporting standards that increase transparency and reproducibility, OHDSI proposed Findable, Accessible, Interoperable, and Reusable (FAIR) phenotype definitions based on APHRODITE. All of the necessary tooling, data models, software and vocabularies are publicly available and released with open-source licenses. [147] As noted in Kashyap et al in evaluating the APHRODITE framework, effective reporting of phenotyping models should include a detailed recipe for data preparation and model training, rather than the pre-trained models themselves, given substantial differences in EHR data across institutions.[115]

Additionally, we observed a lack of rigorous evaluation of phenotyping algorithms, with most studies using standard metrics to evaluate internal validity. We stress further model interrogation for phenotyping, including external validation as well as evaluation of fairness. However, reliable performance evaluation requires a substantial amount of gold-standard labeled data. There is very little work on semi-supervised and weakly-supervised model performance evaluation, and further research is warranted.[148–150]

### Accounting for Misclassified Phenotypes due to Algorithm Errors

As ML phenotyping expands the scope of EHR research, care must be taken when using derived phenotypes for downstream tasks as they are inevitably misclassified due to algorithm errors. In the context of association studies, it is well known in the statistical community that misclassification can lead to diminished statistical power and biased estimation.[151–153] However, statistical methods are often siloed from the informatics community. We advocate for dissemination of existing methods and for methodological developments in “post-phenotyping” inferential and predictive modeling studies.

### Limitations

As the definition of phenotyping is variable within the literature,[12] we used a broad search capturing articles focusing on ML or NLP or phenotyping using EHRs. Following prior work, we limited our scope to select informatics venues.[12,15] Although we have missed articles outside of these journals, our aim is to rigorously characterize the general landscape of ML-based phenotyping, which we believe is captured in the venues considered and in our detailed analyses.

## CONCLUSION

This review summarizes the landscape of ML-based phenotyping between 2018 and 2022. Current literature has laid the groundwork for “deep phenotyping”, but developing standards and methodology for reliable use of a diverse range of phenotypes derived from ML models is necessary for continued EHR-based research.

## Data Availability

All data produced in the present study are available upon reasonable request to the authors

## DATA & CODE AVAILABILITY

The underlying data and R code to replicate our analyses can be found at: https://github.com/jlgrons/ML-EHR-Phenotyping-Review

## ACKNOWLEDGEMENTS

The authors would like to thank Prof. Lei Sun for her useful comments.

## CONTRIBUTIONS

J.G. conceived and designed the study. S.Y. performed the full-text review. J.G. and S.Y. analyzed and interpreted the data. J.G., P.V., and S.Y. drafted and revised the manuscript. J.G., P.V., S.Y., E.S., and K.T. approved the final manuscript.

