## Supplementary Materials for "Machine Learning Approaches for Electronic Health Records Phenotyping: A Methodical Review"

### CONTENT


| | |
|---|---|
| Table S1. Comparison of this article with existing reviews. | **4** |
| Table S2. Database search strategy. | **6** |
| Table S3. Common reasons for article exclusion. | **7** |
| Table S4. List of included papers. | **8** |
| Table S5. List of items recorded from the articles. | **13** |
| Table S6. Common openly available datasets used in the selected articles. | **15** |
| Table S7. Common terminologies used in the selected articles. | **16** |
| Table S8. Common NLP software used in the selected articles. | **17** |
| Table S9. Common data sources to train word embeddings in the selected articles. | **18** |
| Table S10. Common methods to train word embeddings in the selected articles. | **19** |
| Table S11. Variants of deep supervised learning methods in the selected articles. | **20** |
| Table S12. Common prediction performance metrics in the selected articles. | **21** |
| Table S13. Studies comparing machine learning and rule-based approaches. | **22** |
| Figure S1. Weakly-supervised methods compared to rule-based algorithms. | **25** |
| Figure S2. Weakly-supervised methods compared to traditional supervised algorithms. | **26** |
| Figure S3. Traditional supervised methods compared to rule-based algorithms. | **27** |
| Figure S4. Deep supervised methods compared to traditional supervised methods. | **29** |
| Table S14. Studies comparing deep supervised models and traditional supervised models. | **30** |

**Table S1.** Comparison of this article with existing reviews.

| **Authors** | **Years covered** | **PRISMA** | **ML approaches covered** | | | | | **Further directions discussed** |
|---|---|---|---|---|---|---|---|---|
| | | | **TSL** | **DSL** | **SSL** | **WSL** | **USL** | |
| This paper | 2018 - 2022 | ✓ | ✓ | ✓ | ✓ | ✓ | ✓ | Deep phenotyping; Reporting and evaluating standards; Accounting for phenotype error |
| Pendergrass and Crawford [11] | 2010 - 2018 | | | | | | | Semi-supervised or unsupervised methods; Algorithm portability |
| Alzoubi et al.[16] | 2013 - 2017 | | ✓ | ✓ | | | ✓ | Access to a shared EHR database; Natural language processing standards |
| Banda et al.[15] | 2010 - 2017 | | ✓ | | | ✓ | ✓ | Combination of multiple data modalities; Validation of unsupervised phenotype definitions; Deep learning; Collaborative network expansion |
| Zeng et al.[24] | 2010 - 2017 | | ✓ | ✓ | | | ✓ | Data standardization and harmonization; Model generalization and interpretability; Phenotype characterization |
| Robinson et al.[17] | 2010 - 2017 | | ✓ | ✓ | | | ✓ | Data standardization and harmonization; Combination of multiple data modalities |
| McBrien et al.[26] | 2000 - 2016 | ✓ | | | | | | Reporting standards |
| Ford et al.[27] | 2000 - 2015 | | ✓ | | | | | Reporting standards for accuracy; Algorithm portability; Privacy-preserving algorithms |
| Shivade et al.[12] | 2010 - 2012 | | ✓ | | | | | Data standardization; Open-source tool development; Model interpretability |

**Abbreviations**: PRISMA = Preferred reporting items for systematic reviews and meta-analyses. TSL = Traditional supervised learning. DSL = Deep supervised learning. SSL = Semi-supervised learning. WSL = Weakly-supervised learning. USL = Unsupervised learning. EHR = Electronic health record.

**Notes:**

- Pendergrass and Crawford,[11] Zeng et al.,[24] and Robinson et al.[17] did not provide an explicit year range covered by their review. We screened the references and reported the years covered.
- Pendergrass and Crawford [11] performed a general discussion on EHR-based phenotyping, with a focus on rule-based methods. McBrien et al.[26] focused primarily on the evaluation and model performance of existing phenotyping methods.

**Table S2.** Database search strategy.

| **Database** | **Search String** |
|---|---|
| PubMed<br>(n = 745) | ((("machine learning" OR "statistical machine learning" OR "deep learning" OR "unsupervised machine learning" OR "supervised machine learning" OR "semi-supervised machine learning" OR "reinforcement learning" OR "federated learning" OR "transfer learning" OR "distributed learning" OR "NLP" OR "natural language processing" ) OR (machine learning[Mesh] OR natural language processing[Mesh])) OR (("phenotyp*" OR "cohort identification") OR ( phenotype[Mesh] OR cohort analysis[Mesh]))<br>AND (("electronic health records" OR "electronic medical records" OR "EHR" OR "EMR" OR "EHRs" OR "EMRs") OR (electronic health records[Mesh] OR electronic medical records[Mesh])))<br>AND (("2018/01/01"[PDat] : "2022/04/14"[PDat])<br>AND (“Journal of the American Medical Informatics Association: JAMIA”[ta] OR “PloS One”[ta] OR “Journal of Biomedical Informatics”[ta] OR "JAMIA Open"[ta] OR"AMIA Annu Symp Proc")) |
| Web of Science<br>(n = 651) | (TS =((((machine learning) OR (statistical machine learning) OR (deep learning) OR (unsupervised machine learning) OR (supervised machine learning) OR (semi-supervised machine learning) OR (reinforcement learning) OR (federated learning) OR (transfer learning) OR (distributed learning) OR (NLP) OR (natural language processing)) OR ((phenotyp*) OR (cohort identification)))<br>AND ((electronic health records) OR (electronic medical records) OR (EHR) OR (EMR) OR (EHRs) OR (EMRs))))<br>AND (DOP=(2018-01-01/2022-04-14))<br>AND (SO=((Journal of the American Medical Informatics Association) OR (PloS One) OR (Journal of Biomedical Informatics) OR (JAMIA Open) OR (AMIA Annual Symposium proceedings AMIA Symposium)))<br><br>**Search in** - All Databases<br>**Collections** - All |

**Note:** For AMIA papers that have a gap between acceptance date and publish date, we follow the convention of Web of Science and record the acceptance year as criteria.

**Table S3.** Common reasons for article exclusion.

| **Primary reason for exclusion** | **Example** | **Representative paper(s)** |
|---|---|---|
| Did not consider phenotyping | Development of named entity recognition tools to process clinical free-text without an emphasis on phenotyping | Zhao S, Cai Z, Chen H, *et al.* Adversarial training based lattice LSTM for Chinese clinical named entity recognition. *J Biomed Inform* 2019;**99**:103290. |
| | Prediction of a future event (eg. mortality, disease onset) | Mahdavi M, Choubdar H, Zabeh E, *et al*. A machine learning based exploration of COVID-19 mortality risk. *PLoS One* 2021;**16**:e0252384.<br><br>Amrollahi F, Shashikumar SP, Razmi F, *et al.* Contextual Embeddings from Clinical Notes Improves Prediction of Sepsis. *AMIA Annu Symp Proc* 2020;**2020**:197–202. |
| | Statistical analysis of disease of risk factors | Hiramoto S, Asano H, Miyamoto T, *et al*. Risk factors and pharmacotherapy for chemotherapy-induced peripheral neuropathy in paclitaxel-treated female cancer survivors: A retrospective study in Japan. *PLoS One* 2021;**16**:e0261473. |
| | Development of de-identification methods | Liao S, Kiros J, Chen J, *et al.* Improving domain adaptation in de-identification of electronic health records through self-training. *J Am Med Inform Assoc* 2021;**28**:2093–100. |
| Did not use EHRs as the primary data source | Focus on administrative data | Philip G, Djerboua M, Carlone D, *et al*. Validation of a hierarchical algorithm to define chronic liver disease and cirrhosis etiology in administrative healthcare data. *PLoS One* 2020;**15**:e0229218. |
| Did not use machine learning | Use of a rule-based natural language processing method or keyword search | Adekkanattu P, Sholle ET, DeFerio J, *et al.* Ascertaining Depression Severity by Extracting Patient Health Questionnaire-9 (PHQ-9) Scores from Clinical Notes. *AMIA Annu Symp Proc* 2018;**2018**:147–56. |
| | Use of expert-defined rules | Fukasawa T, Takahashi H, Kameyama N, *et al.* Development of an electronic medical record-based algorithm to identify patients with Stevens-Johnson syndrome and toxic epidermal necrolysis in Japan. *PLoS One* 2019;**14**:e0221130. |
| Reviews. editorials, or perspectives | Systematic review | Koleck TA, Dreisbach C, Bourne PE, *et al*. Natural language processing of symptoms documented in free-text narratives of electronic health records: a systematic review. *J Am Med Inform Assoc* 2019;**26**:364–79. |

**Table S4.** List of included papers.

- Afshar M, Joyce C, Oakey A, *et al.* A Computable Phenotype for Acute Respiratory Distress Syndrome Using Natural Language Processing and Machine Learning. *AMIA Annu Symp Proc* 2018;**2018**:157–65.
- Maurits MP, Korsunsky I, Raychaudhuri S, *et al.* A framework for employing longitudinally collected multicenter electronic health records to stratify heterogeneous patient populations on disease history. *J Am Med Inform Assoc* 2022;**29**:761–9.
- Geva A, Liu M, Panickan VA, *et al.* A high-throughput phenotyping algorithm is portable from adult to pediatric populations. *J Am Med Inform Assoc* 2021;**28**:1265–9.
- Zhang L, Ding X, Ma Y, *et al.* A maximum likelihood approach to electronic health record phenotyping using positive and unlabeled patients. *J Am Med Inform Assoc* 2020;**27**:119–26.
- Annapragada AV, Donaruma-Kwoh MM, Annapragada AV, *et al.* A natural language processing and deep learning approach to identify child abuse from pediatric electronic medical records. *PLoS One* 2021;**16**:e0247404.
- Wei Q, Ji Z, Li Z, *et al.* A study of deep learning approaches for medication and adverse drug event extraction from clinical text. *J Am Med Inform Assoc* 2020;**27**:13–21.
- Yu Z, Yang X, Dang C, *et al.* A Study of Social and Behavioral Determinants of Health in Lung Cancer Patients Using Transformers-based Natural Language Processing Models. arXiv preprint arXiv:2108.04949.
- Sanyal J, Rubin D, Banerjee I. A weakly supervised model for the automated detection of adverse events using clinical notes. *J Biomed Inform* 2022;**126**:103969.
- Dai H-J, Su C-H, Wu C-S. Adverse drug event and medication extraction in electronic health records via a cascading architecture with different sequence labeling models and word embeddings. *J Am Med Inform Assoc* 2020;**27**:47–55.
- Obeid JS, Davis M, Turner M, *et al.* An artificial intelligence approach to COVID-19 infection risk assessment in virtual visits: A case report. *J Am Med Inform Assoc* 2020;**27**:1321–5.
- Ju M, Nguyen NTH, Miwa M, *et al.* An ensemble of neural models for nested adverse drug events and medication extraction with subwords. *J Am Med Inform Assoc* 2020;**27**:22–30.
- Fialoke S, Malarstig A, Miller MR, *et al.* Application of Machine Learning Methods to Predict Non-Alcoholic Steatohepatitis (NASH) in Non-Alcoholic Fatty Liver (NAFL) Patients. *AMIA Annu Symp Proc* 2018;**2018**:430–9.
- Murray SG, Avati A, Schmajuk G, *et al.* Automated and flexible identification of complex disease: building a model for systemic lupus erythematosus using noisy labeling. *J Am Med Inform Assoc* 2019;**26**:61–5.
- Ni Y, Bachtel A, Nause K, *et al.* Automated detection of substance use information from electronic health records for a pediatric population. *J Am Med Inform Assoc* 2021;**28**:2116–27.
- Erickson J, Abbott K, Susienka L. Automatic address validation and health record review to identify homeless Social Security disability applicants. *J Biomed Inform* 2018;**82**:41–6.
- Ferté T, Cossin S, Schaeverbeke T, *et al.* Automatic phenotyping of electronical health record: PheVis algorithm. *J Biomed Inform* 2021;**117**:103746.
- Thompson HM, Sharma B, Bhalla S, *et al.* Bias and fairness assessment of a natural language processing opioid misuse classifier: detection and mitigation of electronic health record data disadvantages across racial subgroups. *J Am Med Inform Assoc* 2021;**28**:2393–403.
- Mitra A, Rawat BPS, McManus D, *et al.* Bleeding Entity Recognition in Electronic Health Records: A Comprehensive Analysis of End-to-End Systems. *AMIA Annu Symp Proc* 2020;**2020**:860–9.
- Zhou S, Wang N, Wang L, *et al.* CancerBERT: a cancer domain-specific language model for extracting breast cancer phenotypes from electronic health records. *J Am Med Inform Assoc* 2022;:ocac040.
- Han S, Zhang RF, Shi L, *et al.* Classifying social determinants of health from unstructured electronic health records using deep learning-based natural language processing. *J Biomed Inform* 2022;**127**:103984.

- Bhattacharya M, Jurkovitz C, Shatkay H. Co-occurrence of medical conditions: Exposing patterns through probabilistic topic modeling of snomed codes. *J Biomed Inform* 2018;**82**:31–40.
- Xiong Y, Shi X, Chen S, *et al.* Cohort selection for clinical trials using hierarchical neural network. *J Am Med Inform Assoc* 2019;**26**:1203–8.
- Dai H-J, Wang F-D, Chen C-W, *et al.* Cohort selection for clinical trials using multiple instance learning. *J Biomed Inform* 2020;**107**:103438.
- Gehrmann S, Dernoncourt F, Li Y, *et al.* Comparing deep learning and concept extraction based methods for patient phenotyping from clinical narratives. *PLoS One* 2018;**13**:e0192360.
- Malmasi S, Ge W, Hosomura N, *et al.* Comparing information extraction techniques for low-prevalence concepts: The case of insulin rejection by patients. *J Biomed Inform* 2019;**99**:103306.
- Kulshrestha S, Dligach D, Joyce C, *et al.* Comparison and interpretability of machine learning models to predict severity of chest injury. *JAMIA Open* 2021;**4**:ooab015.
- Nori VS, Hane CA, Sun Y, *et al.* Deep neural network models for identifying incident dementia using claims and EHR datasets. *PLoS One* 2020;**15**:e0236400.
- Ogunyemi OI, Gandhi M, Lee M, *et al.* Detecting diabetic retinopathy through machine learning on electronic health record data from an urban, safety net healthcare system. *JAMIA Open* 2021;**4**:ooab066.
- Cohen AM, Chamberlin S, Deloughery T, *et al.* Detecting rare diseases in electronic health records using machine learning and knowledge engineering: Case study of acute hepatic porphyria. *PLoS One* 2020;**15**:e0235574.
- Zhao J, Zhang Y, Schlueter DJ, *et al.* Detecting time-evolving phenotypic topics via tensor factorization on electronic health records: Cardiovascular disease case study. *J Biomed Inform* 2019;**98**:103270.
- Bucher BT, Shi J, Pettit RJ, *et al.* Determination of Marital Status of Patients from Structured and Unstructured Electronic Healthcare Data. *AMIA Annu Symp Proc* 2019;**2019**:267–74.
- Hong N, Wen A, Stone DJ, *et al.* Developing a FHIR-based EHR phenotyping framework: A case study for identification of patients with obesity and multiple comorbidities from discharge summaries. *J Biomed Inform* 2019;**99**:103310.
- Martin JA, Crane-Droesch A, Lapite FC, *et al.* Development and validation of a prediction model for actionable aspects of frailty in the text of clinicians' encounter notes. *J Am Med Inform Assoc* 2021;**29**:109–19.
- Kashyap M, Seneviratne M, Banda JM, *et al.* Development and validation of phenotype classifiers across multiple sites in the observational health data sciences and informatics network. *J Am Med Inform Assoc* 2020;**27**:877–83.
- Docherty M, Regnier SA, Capkun G, *et al.* Development of a novel machine learning model to predict presence of nonalcoholic steatohepatitis. *J Am Med Inform Assoc* 2021;**28**:1235–41.
- Koola JD, Davis SE, Al-Nimri O, *et al.* Development of an automated phenotyping algorithm for hepatorenal syndrome. *J Biomed Inform* 2018;**80**:87–95.
- Wang L, Lakin J, Riley C, *et al.* Disease Trajectories and End-of-Life Care for Dementias: Latent Topic Modeling and Trend Analysis Using Clinical Notes. *AMIA Annu Symp Proc* 2018;**2018**:1056–65.
- Gao J, Xiao C, Glass LM, *et al.* Dr. Agent: Clinical predictive model via mimicked second opinions. *J Am Med Inform Assoc* 2020;**27**:1084–91.
- Gibson TB, Nguyen MD, Burrell T, *et al.* Electronic phenotyping of health outcomes of interest using a linked claims-electronic health record database: Findings from a machine learning pilot project. *J Am Med Inform Assoc* 2021;**28**:1507–17.
- Yu S, Ma Y, Gronsbell J, *et al.* Enabling phenotypic big data with PheNorm. *J Am Med Inform Assoc* 2018;**25**:54–60.
- Kim Y, Meystre SM. Ensemble method-based extraction of medication and related information from clinical texts. *J Am Med Inform Assoc* 2020;**27**:31–8.
- Mahesri M, Chin K, Kumar A, *et al.* External validation of a claims-based model to predict left ventricular ejection fraction class in patients with heart failure. *PLoS ONE* 2021;**16**:e0252903.

- Eisman AS, Shah NR, Eickhoff C, *et al.* Extracting Angina Symptoms from Clinical Notes Using Pre-Trained Transformer Architectures. *AMIA Annu Symp Proc* 2020;**2020**:412–21.
- Lybarger K, Ostendorf M, Thompson M, *et al.* Extracting COVID-19 diagnoses and symptoms from clinical text: A new annotated corpus and neural event extraction framework. *J Biomed Inform* 2021;**117**:103761.
- Chen L, Gu Y, Ji X, *et al.* Extracting medications and associated adverse drug events using a natural language processing system combining knowledge base and deep learning. *J Am Med Inform Assoc* 2020;**27**:56–64.
- Xie K, Gallagher RS, Conrad EC, *et al.* Extracting seizure frequency from epilepsy clinic notes: a machine reading approach to natural language processing. *J Am Med Inform Assoc* 2022;**29**:873–81.
- Ning W, Chan S, Beam A, *et al.* Feature extraction for phenotyping from semantic and knowledge resources. *J Biomed Inform* 2019;**91**:103122.
- Estiri H, Vasey S, Murphy SN. Generative transfer learning for measuring plausibility of EHR diagnosis records. *J Am Med Inform Assoc* 2021;**28**:559–68.
- Liao KP, Sun J, Cai TA, *et al.* High-throughput multimodal automated phenotyping (MAP) with application to PheWAS. *J Am Med Inform Assoc* 2019;**26**:1255–62.
- Estiri H, Strasser ZH, Murphy SN. High-throughput phenotyping with temporal sequences. *J Am Med Inform Assoc* 2021;**28**:772–81.
- Shen F, Peng S, Fan Y, *et al*. HPO2Vec+: Leveraging heterogeneous knowledge resources to enrich node embeddings for the Human Phenotype Ontology. *J Biomed Inform* 2019;**96**:103246.
- Stemerman R, Arguello J, Brice J, *et al.* Identification of social determinants of health using multi-label classification of electronic health record clinical notes. *JAMIA Open* 2021;**4**:ooaa069.
- Carson NJ, Mullin B, Sanchez MJ, *et al*. Identification of suicidal behavior among psychiatrically hospitalized adolescents using natural language processing and machine learning of electronic health records. *PLoS One* 2019;**14**:e0211116.
- Seneviratne MG, Banda JM, Brooks JD, et al. Identifying Cases of Metastatic Prostate Cancer Using Machine Learning on Electronic Health Records. AMIA Annu Symp Proc 2018;2018:1498–504.
- Yang X, Bian J, Fang R, *et al.* Identifying relations of medications with adverse drug events using recurrent convolutional neural networks and gradient boosting. *J Am Med Inform Assoc* 2020;**27**:65–72.
- Xu Z, Chou J, Zhang XS, *et al.* Identifying sub-phenotypes of acute kidney injury using structured and unstructured electronic health record data with memory networks. *J Biomed Inform* 2020;**102**:103361.
- Grundmeier RW, Xiao R, Ross RK, *et al.* Identifying surgical site infections in electronic health data using predictive models. *J Am Med Inform Assoc* 2018;**25**:1160–6.
- Chen T, Dredze M, Weiner JP, *et al.* Identifying vulnerable older adult populations by contextualizing geriatric syndrome information in clinical notes of electronic health records. *J Am Med Inform Assoc* 2019;**26**:787–95.
- Kronzer VL, Wang L, Liu H, *et al*. Investigating the impact of disease and health record duration on the eMERGE algorithm for rheumatoid arthritis. *J Am Med Inform Assoc* 2020;**27**:601–5.
- Goodwin TR, Harabagiu SM. Learning relevance models for patient cohort retrieval. *JAMIA Open* 2018;**1**:265–75.
- Mullin S, Zola J, Lee R, *et al.* Longitudinal K-means approaches to clustering and analyzing EHR opioid use trajectories for clinical subtypes. *J Biomed Inform* 2021;**122**:103889.
- Gong J, Simon GE, Liu S. Machine learning discovery of longitudinal patterns of depression and suicidal ideation. *PLoS ONE* 2019;**14**:e0222665.
- Badger J, LaRose E, Mayer J, *et al.* Machine learning for phenotyping opioid overdose events. *J Biomed Inform* 2019;**94**:103185.
- Hassanzadeh H, Karimi S, Nguyen A. Matching patients to clinical trials using semantically enriched document representation. *J Biomed Inform* 2020;**105**:103406.
- Chen C-J, Warikoo N, Chang Y-C, *et al*. Medical knowledge infused convolutional neural networks for cohort selection in clinical trials. *J Am Med Inform Assoc* 2019;**26**:1227–36.

- Bejan CA, Angiolillo J, Conway D, *et al.* Mining 100 million notes to find homelessness and adverse childhood experiences: 2 case studies of rare and severe social determinants of health in electronic health records. *J Am Med Inform Assoc* 2018;**25**:61–71.
- Topaz M, Murga L, Gaddis KM, *et al*. Mining fall-related information in clinical notes: Comparison of rule-based and novel word embedding-based machine learning approaches. *J Biomed Inform* 2019;**90**:103103.
- Wu S, Liu S, Sohn S, *et al*. Modeling asynchronous event sequences with RNNs. *J Biomed Inform* 2018;**83**:167–77.
- Afshar M, Phillips A, Karnik N, *et al.* Natural language processing and machine learning to identify alcohol misuse from the electronic health record in trauma patients: development and internal validation. *J Am Med Inform Assoc* 2019;**26**:254–61.
- Meaney C, Escobar M, Moineddin R, *et al.* Non-negative matrix factorization temporal topic models and clinical text data identify COVID-19 pandemic effects on primary healthcare and community health in Toronto, Canada. *J Biomed Inform* 2022;**128**:104034.
- Ibrahim ZM, Wu H, Hamoud A, *et al*. On classifying sepsis heterogeneity in the ICU: insight using machine learning. *J Am Med Inform Assoc* 2020;**27**:7.
- Lyudovyk O, Shen Y, Tatonetti NP, *et al*. Pathway analysis of genomic pathology tests for prognostic cancer subtyping. *J Biomed Inform* 2019;**98**:103286.
- Soni S, Roberts K. Patient Cohort Retrieval using Transformer Language Models. *AMIA Annu Symp Proc* 2020;**2020**:1150–9.
- Zheng NS, Feng Q, Kerchberger VE, *et al.* PheMap: a multi-resource knowledge base for high-throughput phenotyping within electronic health records. *J Am Med Inform Assoc* 2020;**27**:1675–87.
- Henderson J, He H, Malin BA, *et al.* Phenotyping through Semi-Supervised Tensor Factorization (PSST). *AMIA Annu Symp Proc* 2018;**2018**:564–73.
- Sinnott JA, Cai F, Yu S, *et al.* PheProb: probabilistic phenotyping using diagnosis codes to improve power for genetic association studies. *J Am Med Inform Assoc* 2018;**25**:1359–65.
- Choudhury O, Park Y, Salonidis T, *et al.* Predicting Adverse Drug Reactions on Distributed Health Data using Federated Learning. *AMIA Annu Symp Proc* 2019;**2019**:313–22.
- Segura-Bedmar I, Colón-Ruíz C, Tejedor-Alonso MÁ, *et al.* Predicting of anaphylaxis in big data EMR by exploring machine learning approaches. *J Biomed Inform* 2018;**87**:50–9.
- Buckland RS, Hogan JW, Chen ES. Selection of Clinical Text Features for Classifying Suicide Attempts. *AMIA Annu Symp Proc* 2020;**2020**:273–82.
- Cade BE, Hassan SM, Dashti HS, *et al.* Sleep apnea phenotyping and relationship to disease in a large clinical biobank. *JAMIA Open* 2022;**5**:ooab117.
- Ben-Assuli O, Jacobi A, Goldman O, *et al.* Stratifying individuals into non-alcoholic fatty liver disease risk levels using time series machine learning models. *J Biomed Inform* 2022;**126**:103986.
- Hubbard RA, Xu J, Siegel R, *et al.* Studying pediatric health outcomes with electronic health records using Bayesian clustering and trajectory analysis. *J Biomed Inform* 2021;**113**:103654.
- Afshar M, Joyce C, Dligach D, *et al.* Subtypes in patients with opioid misuse: A prognostic enrichment strategy using electronic health record data in hospitalized patients. *PLoS One* 2019;**14**:e0219717.
- Ahuja Y, Zhou D, He Z, *et al.* sureLDA: A multidisease automated phenotyping method for the electronic health record. *J Am Med Inform Assoc* 2020;**27**:1235–43.
- Liu Q, Woo M, Zou X, *et al*. Symptom-based patient stratification in mental illness using clinical notes. *J Biomed Inform* 2019;**98**:103274.
- Kim Y, Lhatoo S, Zhang G-Q, *et al.* Temporal phenotyping for transitional disease progress: An application to epilepsy and Alzheimer’s disease. *J Biomed Inform* 2020;**107**:103462.
- To D, Joyce C, Kulshrestha S, *et al.* The Addition of United States Census-Tract Data Does Not Improve the Prediction of Substance Misuse. *AMIA Annu Symp Proc* 2021;**2021**:1149–58.

- Kreuger AL, Middelburg RA, Beckers EAM, *et al.* The identification of cases of major hemorrhage during hospitalization in patients with acute leukemia using routinely recorded healthcare data. *PLoS One* 2018;**13**:e0200655.
- Ni Y, Alwell K, Moomaw CJ, *et al.* Towards phenotyping stroke: Leveraging data from a large-scale epidemiological study to detect stroke diagnosis. *PLoS One* 2018;**13**:e0192586.
- Apostolova E, Uppal A, Galarraga JE, *et al*. Towards Reliable ARDS Clinical Decision Support: ARDS Patient Analytics with Free-text and Structured EMR Data. *AMIA Annu Symp Proc* 2019;**2019**:228–37.
- Feller DJ, Zucker J, Don't Walk OB, *et al.* Towards the Inference of Social and Behavioral Determinants of Sexual Health: Development of a Gold-Standard Corpus with Semi-Supervised Learning. *AMIA Annu Symp Proc* 2018;**2018**:422–9.
- Lu S, Chen R, Wei W, *et al*. Understanding Heart Failure Patients EHR Clinical Features via SHAP Interpretation of Tree-Based Machine Learning Model Predictions. *AMIA Annu Symp Proc* 2021;**2021**:813–22.
- Wang Y, Zhao Y, Therneau TM, *et al.* Unsupervised machine learning for the discovery of latent disease clusters and patient subgroups using electronic health records. *J Biomed Inform* 2020;**102**:103364.
- Zhou F, Gillespie A, Gligorijevic D, *et al*. Use of disease embedding technique to predict the risk of progression to end-stage renal disease. *J Biomed Inform* 2020;**105**:103409.
- Ling AY, Kurian AW, Caswell-Jin JL, *et al*. Using natural language processing to construct a metastatic breast cancer cohort from linked cancer registry and electronic medical records data. *JAMIA Open* 2019;**2**:528–37.
- Shi J, Liu S, Pruitt LCC, *et al*. Using Natural Language Processing to improve EHR Structured Data-based Surgical Site Infection Surveillance. *AMIA Annu Symp Proc* 2019;**2019**:794–803.
- Chu J, Dong W, He K, *et al*. Using neural attention networks to detect adverse medical events from electronic health records. *J Biomed Inform* 2018;**87**:118–30.
- Lybarger K, Yetisgen M, Ostendorf M. Using Neural Multi-task Learning to Extract Substance Abuse Information from Clinical Notes. *AMIA Annu Symp Proc* 2018;**2018**:1395–404.
- Klann JG, Estiri H, Weber GM, *et al.* Validation of an internationally derived patient severity phenotype to support COVID-19 analytics from electronic health record data. *J Am Med Inform Assoc* 2021;**28**:1411–20.
- Banerjee I, Li K, Seneviratne M, *et al*. Weakly supervised natural language processing for assessing patient-centered outcome following prostate cancer treatment. *JAMIA Open* 2019;**2**:150–9.

**Table S5.** List of items recorded from the articles.

| Aspect | Category | Items recorded | Type(s) |
|---|---|---|---|
| Summary | General information | The online article link, source, PMID, title, authors, journal name, year of publication, and the abstract. | String, Number |
| | Primary goal | 1-2 sentences describing the goal of the paper. | String |
| | Primary contribution | Primary contribution of the paper are recorded:<br>(i) Methods: papers that propose a new method or new architecture of deep learning variants<br>(ii) Application: papers that apply existing methods to a specific task<br>(iii) Comparative studies: papers that compare methods<br>(iv) Evaluation: papers that evaluate the performance of methods across different data sources, or healthcare settings | String |
| Data source used | Data source | The institution or site(s) that the data are extracted from. | List |
| | Multiple institutions | Whether the research is conducted in multiple institutions. | Boolean |
| | Research network | The research network that the institution belongs to, if any. | String |
| | Publicly available data | Whether the data is publicly available. | Boolean |
| | Competition data | The competition that provided the data, if any. | String |
| | Additional data | Additional data sources other than EHRs, if any. | List |
| | Country | The country that the data comes from. | List |
| | Data size | The size of institution data and size of the cohort of interest. | List |
| | Unstructured data | The type of the unstructured data, if any. | List |
| | Unstructured data language | The language of the clinical notes, if any.. | String |
| | Structured data | The type of the structured data, if any. | List |
| | Common data model (CDM) | The common data model used, if any. | String |
| | Terminology | The terminologies used (eg. ICD-10-CM, LOINC, RxNorm), if any. | List |
| Phenotype considered | Phenotypes | Phenotypes considered in the article. | List |
| | Phenotype classification | The phenotypes considered are classified into either (i) binary, (ii) categorical, or (iii) continuous.<br>They are also classified into disease progression, severity, social determinants of health (SODHs), and subtype or | Integer |

| | | | |
|---|---|---|---|
| | | subgroups, if appropriate. | |
| Methods applied | Traditional machine learning (ML) method | The traditional ML method used to develop the phenotyping model, if any. | List |
| | Deep learning (DL) method | The DL method used, if any. | List |
| | DL method subname | The DL subname of (eg. RNN variants, BERT variants) | List |
| | ML Type | Whether the primary approach was (i) Supervised, (ii) Unsupervised, (iii) Semi-supervised, (iv) Weakly-supervised. | String |
| | Labels for training | Methods used to develop labels for training the model. | String |
| | Labels for testing | Methods used to develop labels for testing the model. | String |
| | Training, validation, and testing size | The size of the training, validation, and/or testing data set. | List |
| | Embedding data | The embeddings used, if any. | List |
| | Embedding method | Method used to train embeddings, if any. | List |
| | Imbalance method | The methods used to deal with class-imbalance data, if any. | String |
| | NLP software | The NLP software used for processing text. | List |
| Reporting and evaluation used | Comparison to rule-based | The rule-based methods compared with, if any. | List |
| | Comparison to traditional ML | The traditional ML methods compared with, if the article focuses on a DL approach and provides a comparison. | List |
| | Best performing method | Method with the best performance using the metrics defined in the article, if any. | String |
| | Model performance metrics | The values of the model performance metrics for the best performing model and the comparator model(s), if any. | List |
| | Fairness metrics | The fairness metrics the article used, if any. | List |
| | Fairness attribute | The protected attribute the article examined for fairness evaluation, if any. | List |
| | Reported demographics | Whether the paper reported the demographics of the study population. | Boolean |
| | Open code | Whether the paper released their source codes. The link to the open-source code is also provided. | Boolean/ List |

**Table S6.** Common openly available datasets used in the selected articles.

| Data type | Data source | Number of articles | | | | | |
|---|---|---|---|---|---|---|---|
| | | **TSL** | **DSL** | **SSL** | **WSL** | **USL** | **Total** |
| Competition data (n = 14) | 2018 National NLP Clinical Challenges (n2c2, formerly i2b2) shared task Track 2: Adverse Drug Events and Medication Extraction in EHRs | | 6 | | | | **6** |
| | 2018 National NLP Clinical Challenges (n2c2, formerly i2b2) shared task Track 1: Cohort Selection for Clinical Trials | 1 | 3 | | | | **4** |
| | 2012 Text REtrieval Conference Medical Records Track (TREC) | 1 | 1 | | | | **2** |
| | 2011 Text REtrieval Conference Medical Records Track (TREC) | 1 | 1 | | | | **2** |
| | 2012 Physionet challenge | | 1 | | | | **1** |
| | 2008 Informatics for Integrating Biology and the Bedside (i2b2) NLP Clinical Challenges: Recognizing Obesity and Co-morbidities in Sparse Data | 1 | | | | | **1** |
| Other (n = 15) | MIMIC-III | 1 | 9 | | 2 | 3 | **15** |
| | MTSamples | | 1 | | | | **1** |

**Abbreviations:** TSL = Traditional supervised learning. DSL = Deep supervised learning. SSL = Semi-supervised learning. WSL = Weakly-supervised learning. USL = Unsupervised learning.

**Note**: Some papers used multiple openly available data sources.

**Table S7.** Common terminologies used in the selected articles. A terminology is presented if it is used in more than one article.

| **Terminology** | **Number of articles** | | | | | |
|---|---|---|---|---|---|---|
| | **TSL** | **DSL** | **SSL** | **WSL** | **USL** | **Total** |
| ICD-9 (International Classification of Diseases, Ninth Revision) and ICD-9-CM (International Classification of Diseases, Ninth Revision, Clinical Modification) | 18 | 7 | 4 | 5 | 8 | **42** |
| UMLS (Unified Medical Language System) | 11 | 8 | 1 | 8 | 3 | **31** |
| ICD-10 (International Classification of Diseases, Tenth Revision) | 11 | 4 | 3 | 1 | 1 | **20** |
| SNOMED-CT (Systematized Nomenclature of Medicine, Clinical Terms) | 2 | 4 | | 3 | 3 | **12** |
| RxNorm | 3 | 2 | 1 | 2 | 1 | **9** |
| CPT (Current Procedural Terminology) | 2 | 3 | | 2 | | **7** |
| Phecode | | | 2 | 3 | 2 | **7** |
| ICD (Version unspecified) | | | | 4 | 1 | **5** |
| LOINC (Logical Observation Identifiers Names and Codes) | 3 | | | 1 | | **4** |
| ATC (Anatomical Therapeutic Chemical Classification) | 2 | | | | | **2** |
| NDC (National Drug Code) | 2 | | | | | **2** |

**Abbreviations**: TSL = Traditional supervised learning. DSL = Deep supervised learning. SSL = Semi-supervised learning. WSL = Weakly-supervised learning. USL = Unsupervised learning.

**Note**: Some papers used multiple terminologies.

**Table S8.** Common natural language processing (NLP) software used in the selected articles. An NLP software is presented if it is used in more than one article.

| **NLP software** | **Number of articles** | | | | | |
|---|---|---|---|---|---|---|
| | **TSL** | **DSL** | **SSL** | **WSL** | **USL** | **Total** |
| cTAKES | 8 | 8 | 1 | | 2 | **19** |
| NegEx (Negation detection algorithm) | 3 | | | 2 | 1 | **6** |
| NILE | 1 | | | 5 | | **6** |
| NLTK (Python library) | | 4 | | | 1 | **5** |
| MetaMap | 3 | 1 | | | | **4** |
| Stanford CoreNLP | | 2 | | | | **2** |

**Abbreviations:** TSL = Traditional supervised learning. DSL = Deep supervised learning. SSL = Semi-supervised learning. WSL = Weakly-supervised learning. USL = Unsupervised learning.

**Note:** Some papers articles used multiple NLP software.

**Table S9.** Common data sources to train word embeddings in the selected articles. A data source is presented if it is used in more than one article.

| **Data source** | **Number of articles** |
|---|---|
| Unstructured EHR data | 11 |
| Biomedical literature | 10 |
| MIMIC-III database (for phenotyping with MIMIC-III) | 7 |
| MIMIC-III database (for phenotyping with another data source) | 6 |
| Wikipedia | 6 |
| Structured EHR data | 2 |

**Note:** Some papers used more than one data source.

**Table S10.** Common methods to train word embeddings in the selected articles. A method is presented if it is used in more than one article.

| **Embedding method** | **Number of articles** |
|---|---|
| Word2vec | 19 |
| BERT and BERT variants (BioBERT, Bio+Clinical BERT, RoBERTa) | 12 |
| GloVe | 6 |
| FastText | 2 |
| Not specified | 2 |

**Note:** Some papers used more than one embedding method.

**Table S11.** Variants of deep supervised learning methods in the selected articles.

| DL type | Variants | Number of articles |
|---|---|---|
| Recurrent neural networks (RNNs) | Bidirectional long-short term memory (Bi-LSTM) | 9 |
| | Long-short term memory (LSTM) | 4 |
| | Bi-LSTM with conditional random field (Bi-LSTM-CRF) | 3 |
| | Bidirectional gated recurrent unit (Bi-GRU) | 2 |
| | LSTM-CRF | 1 |
| | GRU | 1 |
| | LSTM-Highway-LSTM | 1 |
| | CNN-Highway-LSTM | 1 |
| | CNN-Bi-LSTM-CRF | 1 |
| | Not specified | 2 |
| Convolutional neural networks (CNNs) | Recurrent convolutional neural networks (RCNN) | 1 |
| | Not specified | 10 |
| BERT and variants | Bidirectional Encoder Representations from Transformers (BERT) | 5 |
| | Robustly optimized BERT approach (RoBERTa) | 2 |
| | BioClinicalBERT | 1 |
| | BioBERT | 1 |
| | CancerBERT | 1 |
| Feed-forward neural networks (FFNNs) | Multi-layer perceptron (MLP) | 1 |
| | Not specified | 2 |

**Note**: Some papers used multiple deep learning types.

**Table S12.** Common prediction performance metrics in the selected articles. A metric is presented if it is used in more than one article.

| **Evaluation metric** | **Number of articles** | | | | |
|---|---|---|---|---|---|
| | **TSL** | **DSL** | **SSL** | **WSL** | **Total** |
| Positive predictive value (PPV) | 23 | 26 | 4 | 8 | **61** |
| Sensitivity | 23 | 25 | 2 | 8 | **58** |
| Area under the receiver operating characteristic curve (AUROC) | 15 | 11 | 5 | 11 | **42** |
| F-score | 9 | 26 | | 7 | **42** |
| Specificity | 11 | 6 | | 2 | **19** |
| Accuracy | 8 | 4 | | 5 | **17** |
| Negative predictive value (NPV) | 7 | 1 | 2 | 5 | **15** |
| Area under the precision-recall curve (AUPRC) | 2 | 4 | | 2 | **8** |
| Calibration plot | 3 | 2 | | | **5** |
| Log loss | 1 | 1 | 1 | | **3** |
| Brier score | 1 | 1 | | | **2** |
| Hamming loss | | 2 | | | **2** |
| Matthews correlation coefficient | 1 | 1 | | | **2** |
| Normalized discounted cumulative gain | 1 | 1 | | | **2** |

**Abbreviations:** TSL = Traditional supervised learning. DSL = Deep supervised learning. SSL = Semi-supervised learning. WSL = Weakly-supervised learning.

**Note**: Most papers reported multiple metrics.

**Table S13.** Studies comparing machine learning (ML) and rule-based approaches.

| ML type | ML-based method | Phenotype | Gold standard | Used Structured data | Used Unstructured data | Rule-based method |
|---|---|---|---|---|---|---|
| Traditional supervised (n = 10) | Support vector machine [155] | Acute Respiratory Distress Syndrome | Chart review | | ✓ | Keyword search |
| | Logistic regression [46] | Alcohol abuse | AUDIT Questionnaire | | ✓ | Keyword search |
| | Logistic regression<br>Support vector machine<br>Decision trees<br>Random forest [156] | Obesity and multiple comorbidities | Chart review | | ✓ | Rule-based algorithm |
| | Support vector machine [158] | Clinical trial eligibility for n2c2 2018 challenge | Chart review | | ✓ | Self-designed rule-based algorithm |
| | Logistic regression with Gradient Boosting [136] | COVID-19 | Chart review | ✓ | | Diagnosis codes<br>Procedure codes<br>Medications<br>Lab results |
| | L1 penalized logistic regression [154] | Hepatorenal Syndrome | Chart review | ✓ | ✓ | Disease-related ICD codes |
| | Random forest [157] | Marital status | Chart review | ✓ | ✓ | Keyword search enriched by UMLS |
| | Random forest [37] | Metastatic Prostate Cancer | Chart review | ✓ | | Disease-related ICD codes |
| | Super Learner [35] | Rhabdomyolysis | Lab results | ✓ | | Disease-related ICD codes |
| | Random forest [74] | Stroke | Chart review | ✓ | | Disease-related ICD codes |
| Deep supervised (n = 2) | Bi-LSTM [54] | Alcohol abuse<br>Marijuana abuse<br>Opioid abuse<br>Tobacco abuse | Chart review | ✓ | ✓ | Logic-based rule matcher |

| | | | | | | |
|---|---|---|---|---|---|---|
| | Bi-LSTM<br>Bi-GRU [137] | Insulin resistance | Chart review | | ✓ | Rulebased detection using Canary platform |
| Semi-supervised (n = 1) | PheCAP [100] | Sleep apnea | Chart review | ✓ | ✓ | Disease-related Phecodes |
| Weakly-supervised (n= 8) | Random forest [115] | Appendicitis<br>Type 2 diabetes mellitus<br>Cataracts<br>Heart failure<br>Abdominal aortic aneurysm<br>Epilepsy<br>Peripheral arterial disease<br>Obesity<br>Glaucoma<br>Venous thromboembolism | Rule-based algorithm | ✓ | | Disease-related SNOMED codes |
| | MAP [109] | Asthma<br>Bipolar disorder<br>Schizophrenia<br>Breast cancer<br>Chronic obstructive pulmonary disease<br>Congestive heart failure<br>Coronary artery disease<br>Hypertension<br>Depression<br>Epilepsy<br>Multiple sclerosis<br>Rheumatoid arthritis<br>Type 1 diabetes mellitus<br>Type 2 diabetes mellitus<br>Crohn's disease<br>Ulcerative colitis | Chart review | ✓ | ✓ | Disease-related ICD codes |
| | Random forest [118] | Fall | Chart review | | ✓ | Self-designed rule-based algorithm |
| | L2 logistic regression [38] | Metastatic breast cancer | Chart review | ✓ | ✓ | Disease-related ICD codes |

| | | | | | | |
|---|---|---|---|---|---|---|
| | PheProb [111] | Rheumatoid arthritis | Chart review | ✓ | | Disease-related Phecodes |
| | PheNorm [108] | Rheumatoid arthritis<br>Crohn's disease<br>Ulcerative colitis<br>Coronary artery disease | Chart review | ✓ | ✓ | Disease-related ICD codes |
| | PheMAP [110] | Type 2 diabetes mellitus Dementia Hypothyroidism | Chart review | ✓ | ✓ | Disease-related Phecodes |
| | Multinomial logistic regression [119] | Urinary incontinence; Bowel dysfunction | Chart review | | ✓ | Disease-related ICD codes |

**Figure S1.** Phenotyping algorithm performance of weakly-supervised methods compared to rule-based algorithms. Metrics are reported as percentages. PPV = positive predictive value. NPV = negative predictive value. AUROC = area under the receiver operating characteristic curve.

**(a)** Weakly-supervised learning outperforms rule-based approach across all reported metrics.

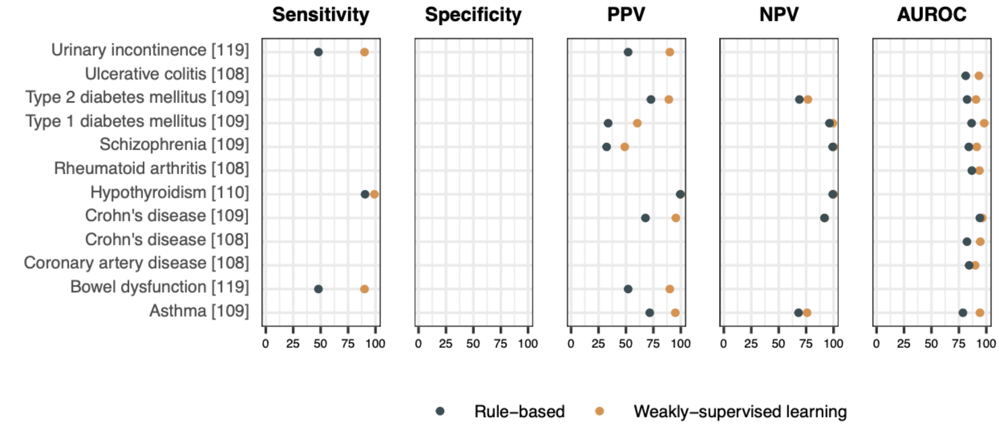


**(b)** Weakly-supervised learning outperforms rule-based approach with respect to PPV.

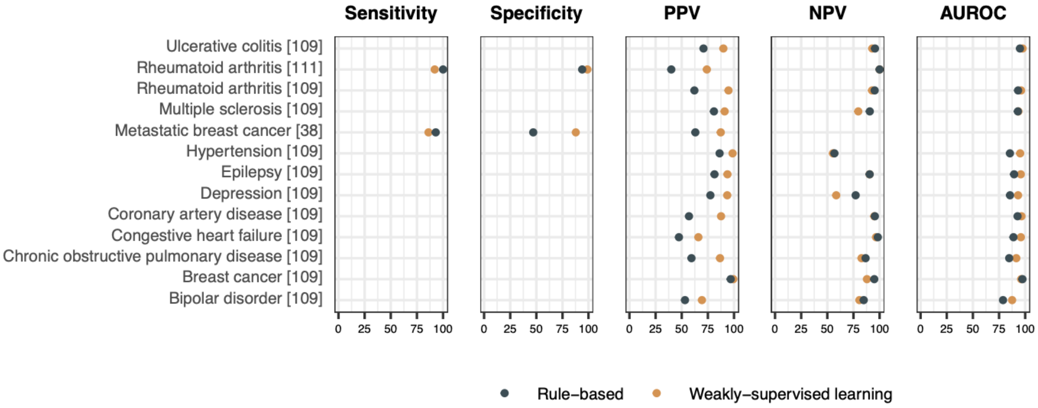


**(c)** Weakly-supervised learning outperforms rule-based approach with respect to sensitivity**.**

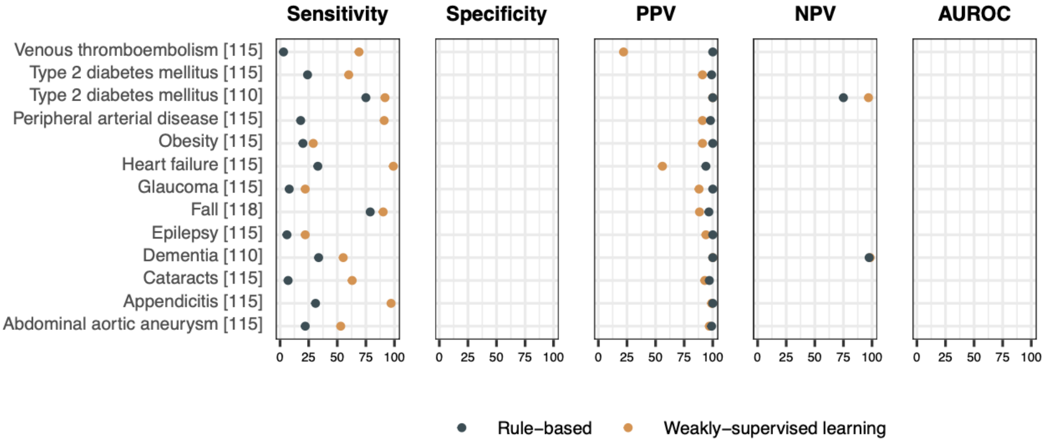

**Figure S2.** Phenotyping algorithm performance of weakly-supervised methods compared to traditional supervised algorithms. Metrics are reported as percentages. Only area under the receiver operating characteristic curve (AUROC) was reported in the articles.

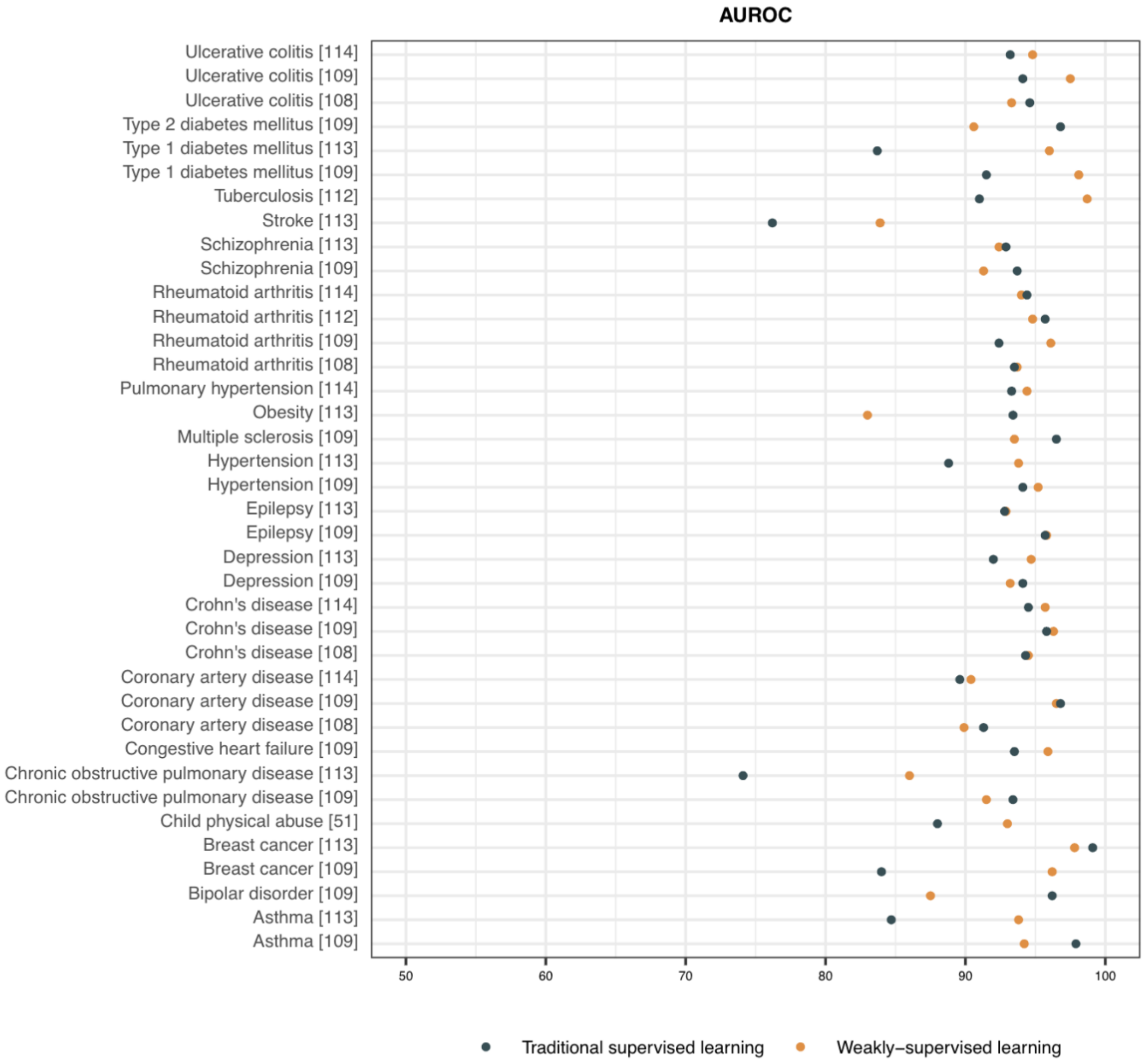

**Figure S3.** Phenotyping algorithm performance of traditional supervised methods compared to rule-based algorithms. Metrics are reported as percentages. PPV = positive predictive value. NPV = negative predictive value. AUROC = area under the receiver operating characteristic curve.

**(a)** Traditional supervised learning outperforms rule-based approach across all reported metrics.

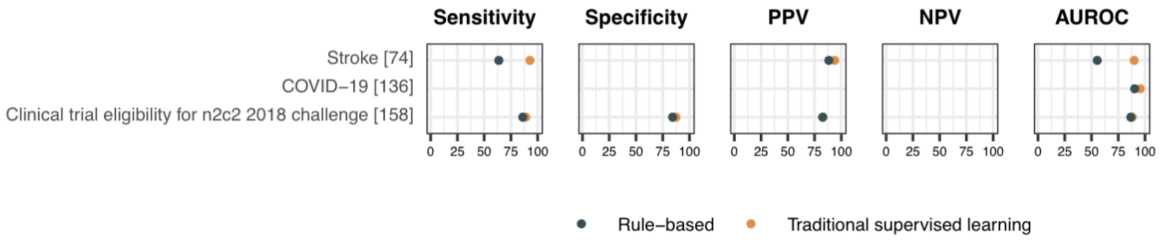


**Note**: [158] reported the micro-averaged metrics for predicting 13 clinical trial selection criteria.

**(b)** Traditional supervised learning outperforms rule-based approach with respect to PPV.

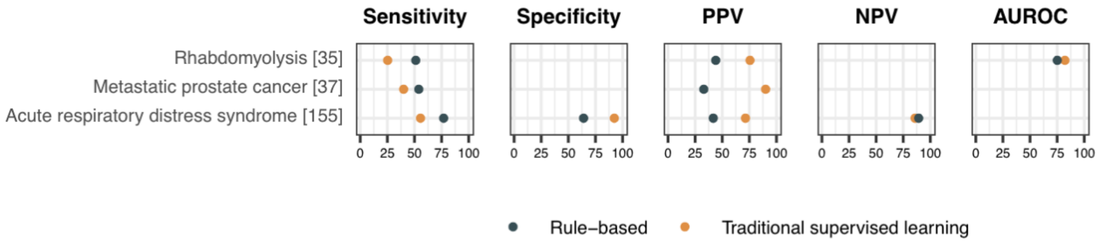


**(c)** Traditional supervised learning outperforms rule-based approach with respect to sensitivity.

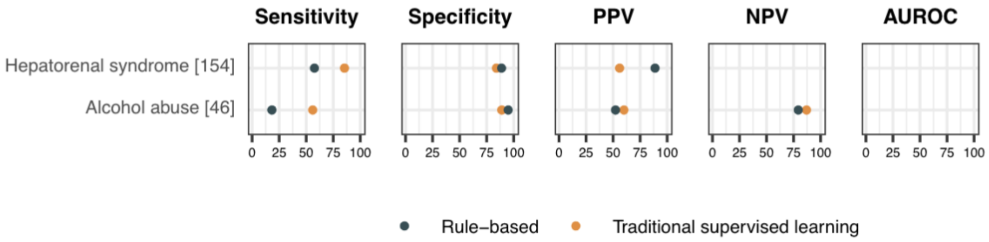


**(d)** Rule-based approach outperforms traditional supervised learning.

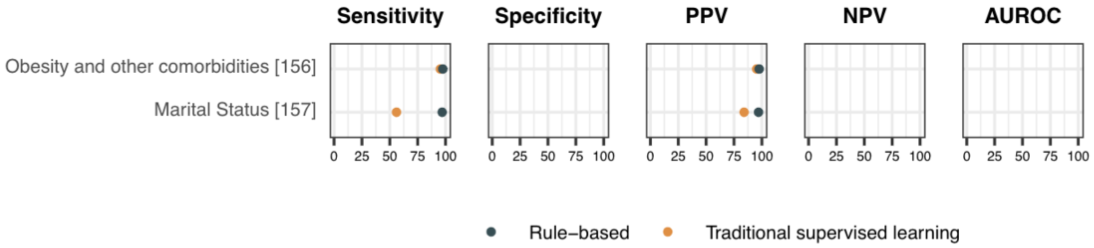


**Note**: [156] reported the micro-averaged metrics for predicting asthma, atherosclerotic cardiovascular disease, congestive heart failure, depression, diabetes mellitus, gallstones/cholecystectomy, gastroesophageal reflux disease, gout, hypercholesterolemia, hypertension, hypertriglyceridemia, obstructive sleep apnea, osteoarthritis, peripheral vascular disease, venous insufficiency.

**Figure S4.** Phenotyping algorithm performance of deep supervised methods compared to traditional supervised methods. Metrics are reported as percentages. AUROC = area under the receiver operating characteristic curve. None of the deep learning papers reported a negative predictive value. A few articles only reported the F-score, which are not included in this figure.

**(a)** Deep supervised learning outperforms traditional supervised learning across all reported metrics.

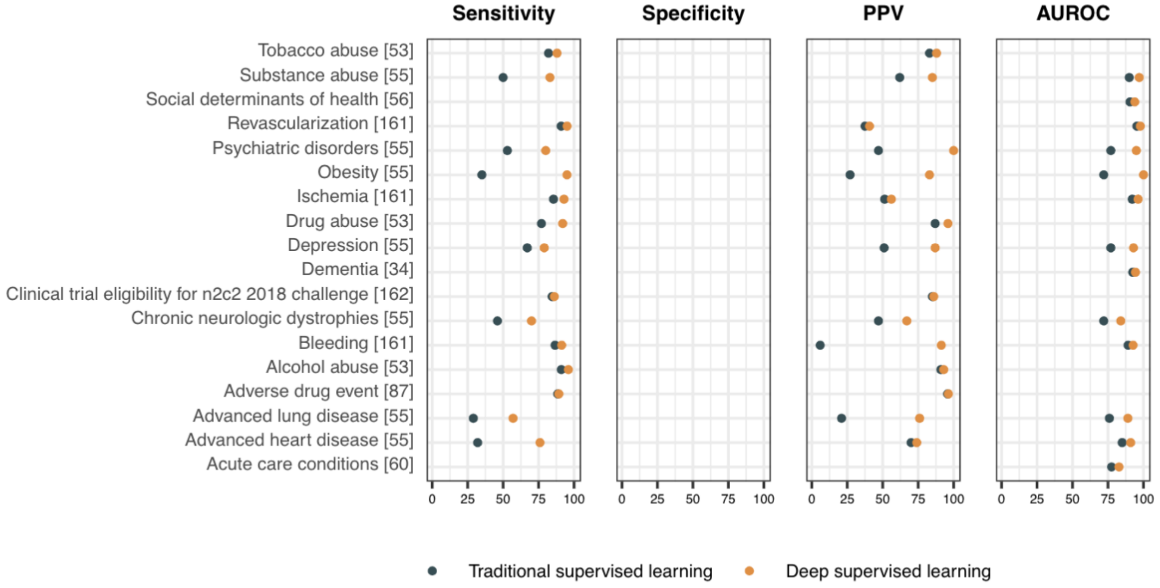


**Note**: [53] reported the micro-averaged metrics for predicting the status of unknown, none, current, and past alcohol abuse, drug abuse, and tobacco abuse. [56] reported the average AUROC for predicting the social determinants of health including homelessness, food stamps, employment, financial resource, insurance status, and social support. [162] reported the micro-averaged metrics for the 13 clinical trial criteria prediction. [60] predicted 25 chronic or critical conditions and only reported averaged values such as micro/macro/weighted AUROC. The micro-AUROC is reported in the plot.

**(b)** Deep supervised learning outperforms traditional supervised learning with respect to PPV.

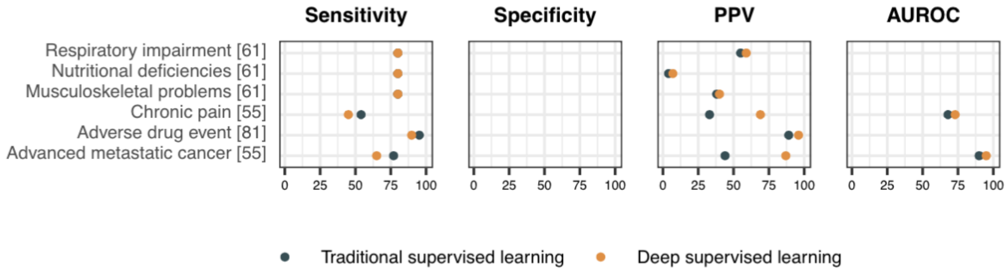

**(c)** Deep supervised learning outperforms traditional supervised learning with respect to sensitivity.

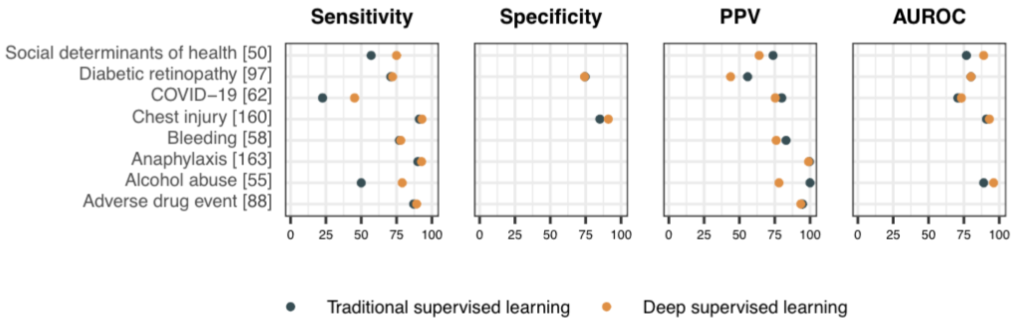


**Note**: [50] reported the micro-averaged metrics for predicting social determinants of health including financial resource, education, insurance status, homelessness, iteraction with the legal system, employment, sexual orientation, social support, spiritual life, tobacco abuse, support circumstances and networks, transportation, internet or cellphone use.

**(d)** Traditional supervised learning outperforms deep supervised learning.

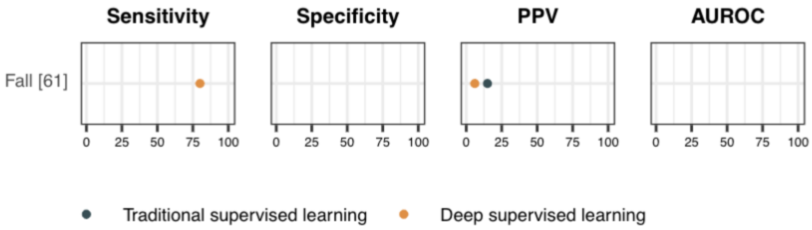

**Table S14.** Studies comparing deep supervised models and traditional supervised models.

| Deep learning type | Deep learning methods | Phenotype | Gold standard | Used structured data | Used unstructured data | Traditional supervised methods |
|---|---|---|---|---|---|---|
| RNN (n = 8) | Bi-LSTM-CRF [80] | Adverse drug event | Chart review | | ✓ | Conditional random field (CRF) |
| | Bi-LSTM-CRF [81] | Adverse drug event | Chart review | | ✓ | Conditional random field (CRF) |
| | Bi-LSTM [87] | Adverse drug event | Chart review | | ✓ | Conditional random field (CRF) |
| | Bi-LSTM-CRF<br>CNN-Bi-LSTM-CRF [88] | Adverse drug event | Chart review | | ✓ | Conditional random field (CRF) |
| | RNN [61] | (Aspects of frailty)<br>Musculoskeletal problems<br>Nutritional deficiencies<br>Respiratory impairment<br>Fall | Chart review | ✓ | ✓ | Elastic-net penalized<br>Logistic regression<br>Random forest |
| | RNN [60] | Acute kidney injury<br>Acute stroke<br>Acute myocardial infarction<br>Cardiac dysrhythmias<br>Chronic kidney disease<br>Chronic obstructive pulmonary disease<br>Complications of surgical/medical care<br>Conduction disorders<br>Congestive heart failure<br>Coronary artery disease<br>Diabetes mellitus with complications;<br>Diabetes mellitus without complication<br>Disorders of lipid metabolism<br>Essential hypertension | ICD codes | ✓ | | L2-penalized logistic regression |

| | | | | | | |
|---|---|---|---|---|---|---|
| | | Fluid and electrolyte disorders<br>Hemorrhage<br>Hypertension with complications<br>Liver disease<br>Lower respiratory disease<br>Upper respiratory disease<br>Pleurisy<br>Pneumonia acute<br>Respiratory impairment<br>Sepsis<br>Shock | | | | |
| | Bi-LSTM [56] | Homelessness<br>Food stamps<br>Employment<br>Financial resource<br>Insurance status<br>Social support | Chart review | | ✓ | Support vector machine (SVM)<br>K-nearest neighbors (KNN)<br>Random forest<br>XGBoost |
| | Bi-LSTM<br>Bi-GRU [137] | Insulin resistance | Chart review | | ✓ | Naive Bayes<br>L2-penalized logistic regression<br>Support vector machine (SVM)<br>Conditional random field (CRF) |
| CNN<br>(n = 5) | CNN [160] | Chest injury | Registry | | ✓ | Elastic-net penalized logistic regression<br>Extreme gradient boosted (XGB) machine |
| | CNN [163] | Anaphylaxis | Chart review | | ✓ | L2-penalized logistic regression<br>Random forest |
| | CNN [62] | COVID-19 | Lab results | ✓ | ✓ | Logistic regression |
| | CNN [162] | Clinical trial eligibility for n2c2 2018 challenge | Chart review | | ✓ | Naive Bayes<br>Support vector machine (SVM)<br>Extreme gradient boosted (XGB) machine |

| | | | | | | |
|---|---|---|---|---|---|---|
| | CNN [55] | Metastatic cancer<br>Adverse heart disease<br>Adverse lung disease<br>Chronic neurologic dystrophies<br>Chronic pain<br>Alcohol abuse<br>Tobacco abuse<br>Obesity<br>Psychiatric disorders<br>Depression | Chart review | | ✓ | Logistic regression |
| BERT<br>(n = 1) | BioBERT [58] | Bleeding | Chart review | ✓ | ✓ | Conditional random field (CRF) |
| FFNN<br>(n = 1) | FFNN [97] | Diabetic retinopathy | ICD codes | ✓ | | Random forest;<br>Support vector machine (SVM)<br>XGBoost<br>Ensemble of 4 stacked classifier including random forest and gradient boosting |
| Multiple types of deep learning neural networks<br>(n = 5) | Bi-LSTM<br>CNN [161] | Ischemia<br>Revascularization<br>Bleeding | Chart review | | ✓ | L2-penalized logistic regression Random forest |
| | Bi-LSTM<br>FFNN [34] | Dementia | Rule-based algorithm | ✓ | ✓ | Boosted trees |
| | LSTM<br>CNN<br>BERT [50] | Financial resource<br>Education<br>Insurance status<br>Homelessness<br>Interaction with the legal system<br>Employment<br>Sexual orientation<br>Social support<br>Spiritual life<br>Tobacco abuse<br>Support circumstances and networks<br>Transportation<br>Internet or cell phone use | Chart review | | ✓ | L2-penalized logistic regression Random forest |
| | Bi-LSTM<br>CNN [53] | Alcohol abuse<br>Drug abuse<br>Tobacco abuse | Chart review | | ✓ | Maximum entropy |

| | | | | | | |
|---|---|---|---|---|---|---|
| | Bi-LSTM<br>Bi-GRU<br>LSTM<br>GRU<br>CNN<br>MLP [159] | Clinical trial eligibility for n2c2 2018 challenge | Chart review | | ✓ | Support vector machine (SVM)<br>Random forest<br>Logistic regression |
